# Mapping spatial colleague connectivity patterns from individual-level registry data to inform regional pandemic interventions

**DOI:** 10.64898/2026.02.19.26346499

**Authors:** PingPing Song, Sake J. de Vlas, Tom Emery, Luc E. Coffeng

**Affiliations:** Department of Public Health, Erasmus MC, University Medical Center Rotterdam, Rotterdam, The Netherlands; Pandemic and Disaster Preparedness Center, Delft, Rotterdam, The Netherlands; Department of Public Administration and Sociology, Erasmus University Rotterdam, Rotterdam, The Netherlands

## Abstract

A concern in infectious disease modelling is how accurately population mixing is incorporated, as it shapes the type and frequency of contacts through which infection spreads, and consequently, estimated intervention effectiveness. Although synthesizing mixing patterns from diary-based surveys is an established framework, geographical information is poorly or sparsely captured. Here we propose a generalizable workflow to quantify geographical connectivity from job registry data covering over 8 million Dutch working population. The derived colleague connectedness shows heterogeneous spatial patterns, quantified from the number of connections per municipality triplet, two residential municipalities and one shared workplace municipality. We demonstrate the utility of this spatial connectivity in signalling regions with elevated outbreak risks. Using SARS-CoV-2 Omicron as an example: a ten-fold increase in within-province connections is associated with a 12-day earlier (95% CI: 2 to 22 days) Omicron onset, and between-province connections associated with an 8-day earlier (95% CI: - 4 to 21 days) onset. These results suggest that the impact of regional interventions shifting spatial connectivity patterns should be expected to vary by region and type of intervention. Together, our findings draw attention of using this highly fine-grained spatial connectivity to enable more regionally tailored and network-targeted policy measures.

## 1. Introduction

Respiratory infectious disease outbreaks such as the SARS-CoV-2 pandemic can unfold unevenly across regions^1,2^. This heterogeneity in regional outbreak dynamics is partly due to differences in the degree of spatially structured contacts through which viruses are transmitted across regions^3^. Within the wider population contact networks, colleague networks play a central role in shaping cross-regional population mixing due to their relatively higher volume compared to friend and family links and wider spatial reach compared to classmate and household connections^4^.

While some large diary-based studies, such as POLYMOD^5–8^ and CoMix^9^, quantify colleague connectivity by the frequency and duration of workplace contacts, they do not record the residential or contact locations of the involved individuals. Transmission models based on these surveys either assume homogeneous workplace mixing^10^ or at most stratified by age groups^11–13^. The lack of spatial heterogeneity in transmission models leads to overestimation of epidemic growth rate^10,14,15^, underestimation of intervention effectiveness^14^, and limits the ability of the models to support regional tailored outbreak responses^16–18^. Alternatively, mobility data based on mobile phone signals offer geographical context^3,18,19^. However, mobile phone signals gathered by commercial data providers are subject to uneven regional representativeness due to differential subscription rates and smartphone use across regions.

To bridge this gap, our study leverages large-scale administrative data from Statistics Netherlands (CBS) to quantify the number of employee pairs working for the same company or even local branch for each year from 2019 to 2022. We chose this data source for its population-scale coverage (over 8 million working individuals in the Netherlands) and high-quality and accurate registry (based on standardized payroll tax registrations). The derived data product of geographical connectivity is quantified by counting the undirected colleague pairs indexed by municipality triplets: two residential municipalities indicating where colleagues live and one joint work municipality indicating where they work. We then aggregated these counts at the municipality level to connectivity patterns at two higher administrative units (safety region and province), at which regional pandemic responses can be implemented in the Netherlands^20–22^. Building on these connectivity patterns, we assessed their epidemiological relevance by applying Bayesian inference to demonstrate associations between colleague connectedness and provincial onset timing of SARS-CoV-2 Omicron emergence. To showcase the utility of our derived data product for supporting region-tailored interventions, we quantified how a regional lockdown and cross-regional travel ban would reduce total colleague links.

## 2. Results

### 2.1. Geographical colleague connectivity patterns

The number of individuals working in the Netherlands with valid payroll tax registrations ranges from 8.0 million in year 2019 to 8.5 million in 2022. We paired colleagues working for the same company branch and counted undirected colleague links per municipality triplet. The colleague link counts are denoted as 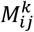, with *i* and *j* being the two colleagues’ residential municipalities and *k* the municipality of their shared workplace. We specifically track workplace location because regional pandemic measures can be applied in workplaces without necessarily affecting areas of residence. To reflect that employees in very large companies are unlikely to have contact with all coworkers, we impose a degree cap, limiting the maximum number of colleagues each individual is allowed to have. With a cap being 100, there were 132 million colleague connections in the year 2021, and a working individual in the Netherlands has, on average, 32 colleague connections. The following results are all for the year 2021 and are based on a cap of 100 colleagues per person.

Figure 1 to Figure 3 visualize the structured patterns of geographical colleague connectivity across the Netherlands, by aggregating colleague counts per municipality triplet 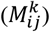 to higher administrative levels. In Figure 1, we distinguish province-level colleague connectivity in two forms: residence-workplace connectivity (Figure 1a) and residence-residence connectivity (Figure 1b). This distinguishment is available due to the triplet location structure in 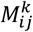. In Figure 1a, grey ribbons dominate, indicating that most workers live and work in the same province. Still, coloured ribbons show substantial cross-province commuting. For example, from the perspective of the origin provinces (top half), many Gelderland (GD) residents work in Utrecht (UT), and Utrecht (UT) residents work in Noord-Holland (NH). From the perspective of the destination provinces (bottom half), Gelderland (GD), Utrecht (UT), Noord-Holland (NH), Zuid-Holland (ZH) and Noord-Brabant (NB) are destinations of notably more cross-province colleague connections than other provinces.

**Figure 1.**
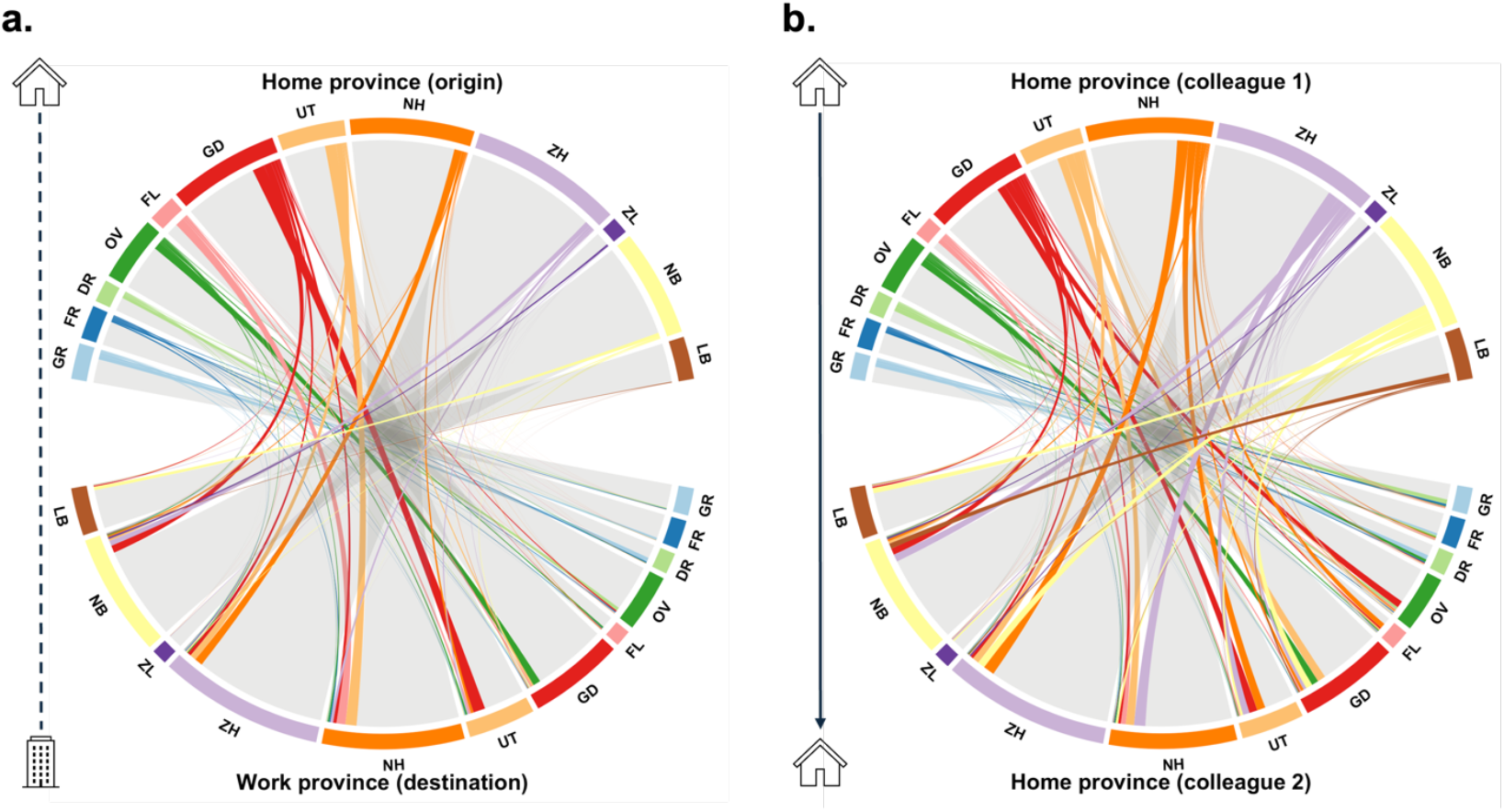
Colleague links between provinces grouped by home-work or home-home pairs. Each panel visualizes the volume of colleague connections (in chords) between pairs of Dutch provinces. Grey ribbons denote within-province connections, while coloured ribbons indicate cross-province links. **(a)** Colleague connectivity between residential and workplace provinces. The top represents the residential province, and the bottom half represents where that individual works, indicating where individuals potentially commute for work. **(b)** Colleague connectivity between two residential provinces. The top half represents the residential province of one colleague (ego), and the bottom half represents the residential province of the colleague working in the same workplace (alter).

Spatial distribution of colleague connections by residential province pairs is visualized in Figure 1b. Similarly, grey ribbons dominate, indicating that most colleagues live in the same province. Even so, notable interprovincial connections displayed with coloured ribbons are non-negligible. For instance, many residents of Zuid-Holland (ZH) share workplaces with those from Noord-Holland (NH), Noord-Brabant (NB) and Utrecht (UT). A structural difference between two panels is that the top and bottom margins in Figure 1b have the same totals per province and the same ribbon width per province pair, due to mirror symmetry of the bidirectional home-home connections. However, the top (home) and bottom (work) halves in Figure 1a have different totals per province and the segments for the same ribbon do not necessarily match in width.

Regional variation in residential province connectedness is further clarified in Figure 2 by differentiating total colleague link volume (Figure 2a) from how links are distributed across province pairs after row-normalization (Figure 2c). Figure 2a mirrors the symmetric structure of Figure 1b and shows that most colleague pairs live within the sane province. Normalization removes differences largely driven by provincial labour force sizes (as summarized in Figure 2b). For transmission models, Figure 2b serves as an input describing the scale of workplace mixing by province, whereas Figure 2c provides the relative distribution of mixing across provinces. The raw data used to generate this figure are provided in supplementary Table S1.

**Figure 2.**
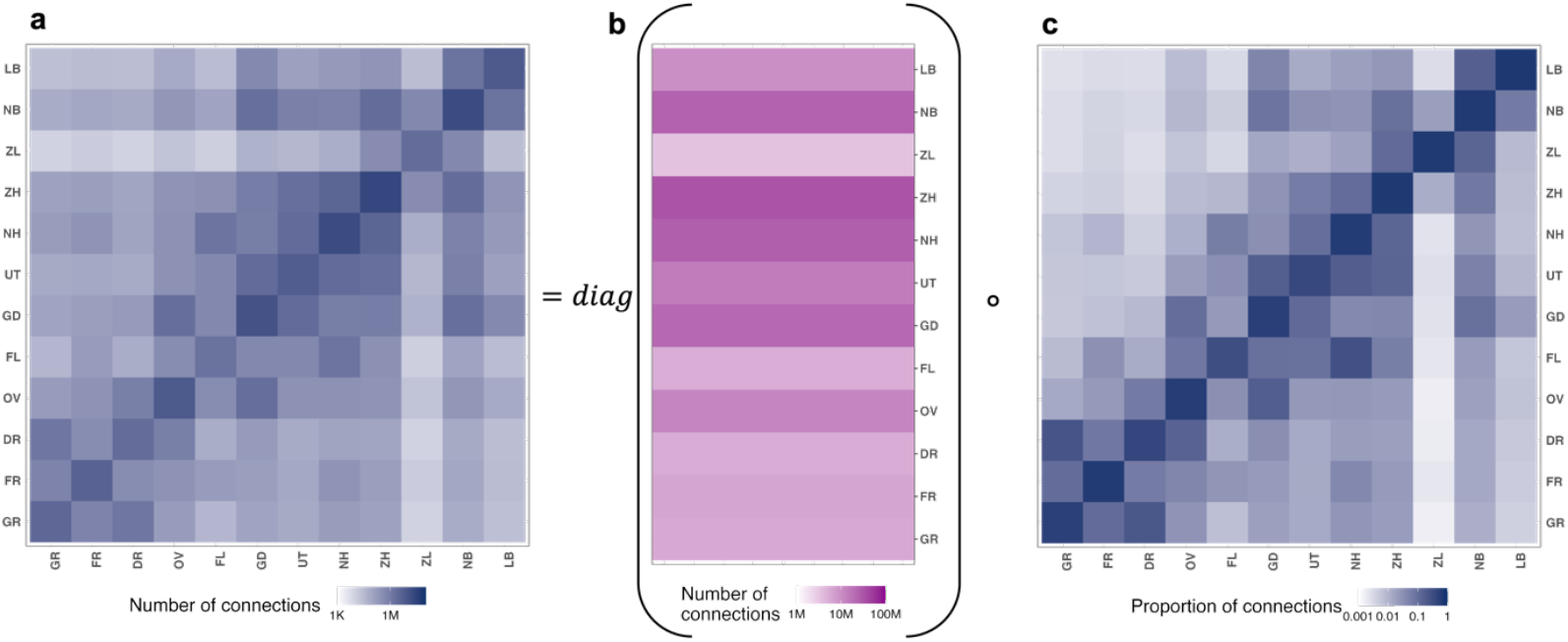
Heterogeneous regional colleague connectivity by residential provinces. **(a)** A matrix with each element being the absolute number of colleague connections between residential province pairs. **(b)** A vector consisting total number of colleague connections per residential province. **(c)** A matrix containing the row-normalized connections between residential province pairs, showing the relative distribution of connections from each province (i.e., the cells in each row add up to one).

Figure 3 subsequently quantifies the heterogeneity of geographical colleague connectivity across administrative scales, reporting for each residential province the share of connections within the same municipality (green), between municipalities in the same province (orange), and between provinces (blue). The share of within-municipality connections is similar across provinces, ranging from 16% in Utrecht (UT) to 29% in Zeeland (ZL). In contrast, there is more variation in within-province and between-province connections. Flevoland (FL) has the lowest share of within-province connections (9%) and the highest share of between-province connections (68%), whereas Limburg (LB) shows the opposite pattern, with the highest within-province share (49%) and the lowest between-province share (29%). Utrecht (UT) and Drenthe (DR) also display a high share of inter-province connections, though likely for different reasons: Utrecht (UT) functions as a business hub, attracting commuters for institutions such as national banks, whereas residents of Drenthe (DR) are more likely to travel outside the province for work due to limited local employers. Raw data of this figure are provided in supplementary Table S2.

**Figure 3.**
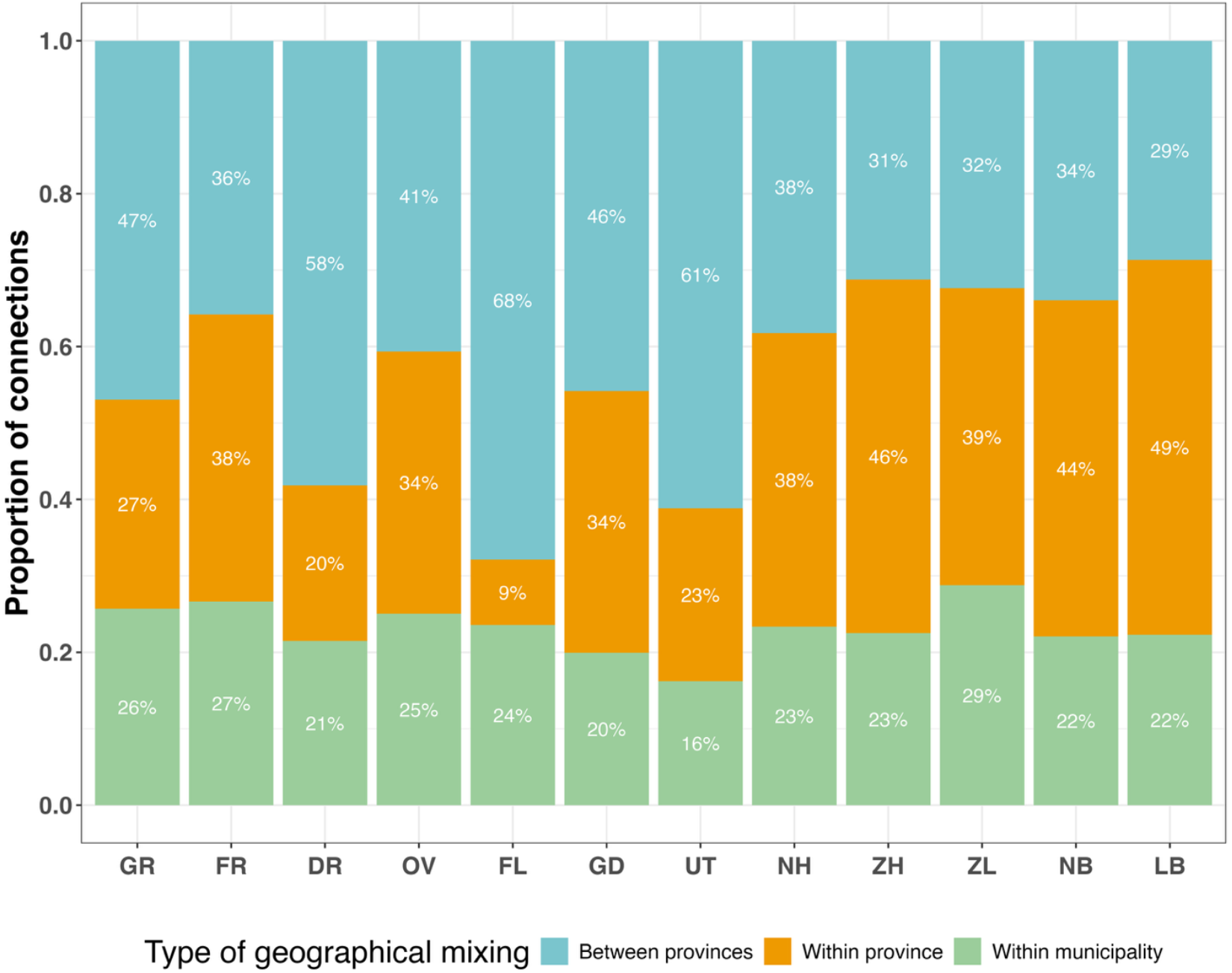
Heterogeneous connectivity shares at different geographic scales. Stacked bar plot shows the proportion of colleague connections originating from individuals living within the same municipality (green), different municipalities within the same province (orange), and between different provinces (blue). Labels indicate the percentage for each connection type per residential province.

### 2.2. From connectivity to Omicron onset across provinces

#### Provincial Omicron onset timing

We estimated the onset timing of Omicron emergence per Dutch province, defined as the first date when Omicron incidence exceeded a prespecified case threshold *I*. As a result, we found that the earliest date at which Omicron reached *I* = 30 cases per week differs across the twelve Dutch provinces by almost one month (shown in supplementary Figure S3). Noord-Holland showed the earliest onset, with a mean date of 26^th^ October 2021 and a 95% credible interval from 23^rd^ to 28^th^ October. After Noord-Holland, the next earliest provinces were Gelderland, Zuid-Holland, Utrecht, and Limburg, with average onsets between 7^th^ and 14^th^ November. Much later onsets were observed in the northern and less populated provinces such as Groningen, Fryslân, Drenthe, and Flevoland, between 20^th^ and 28^th^ November. Uncertainty (shown as horizontal bars) varies, with the widest credible intervals for the least populated provinces Zeeland and Flevoland. Sensitivity analyses of the case threshold *I* (supplementary Figure S4.2) showed that the ranking of onset times is stable across thresholds, with Noord-Holland – home to the country’s largest international airport – consistently estimated as the earliest province to experience the start of the Omicron wave.

#### Associations between connectivity and onsets

We demonstrated to what extent provincial Omicron onset is associated with colleague connectivity patterns per province except Noord-Holland (NH), where first few Omicron cases were likely driven by international transmissions. A negative slope of −0.62 on the standardized natural logarithmic scale (Figure 4) indicates that a tenfold increase in within-province colleague connections is associated with an Omicron onset that is 12 days earlier (95% CI: 2 to 21 days earlier). Similarly, the between-province connectivity slope of −0.36 suggests that a tenfold increase in inter-province colleague connections is associated with an Omicron onset that is 8 days earlier (95% CI: −4 to 20 days earlier).

**Figure 4.**
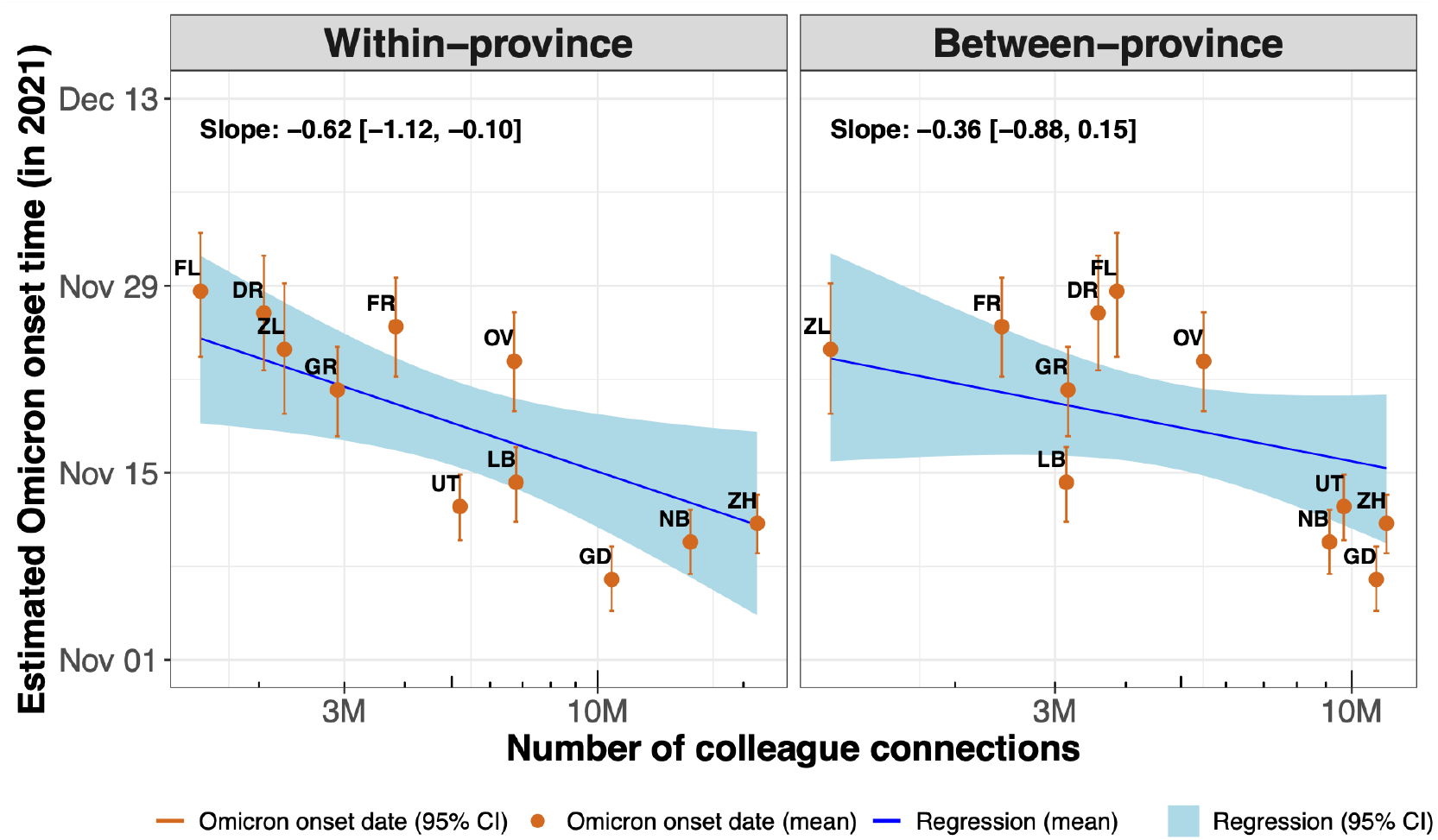
Associations between regional colleague connectivity and Omicron onset timing. Each panel shows estimated Omicron onset time per province (posterior means as circles and 95% credible intervals as error bars) plotted against the number of within- or between-province colleague connections. Blue regression lines and shaded ribbons represent the posterior mean and 95% credible interval from the Bayesian regression inference. The negative trends indicate that provinces with more intra- and inter-province colleague connections are associated with earlier Omicron onset.

In both panels of Figure 4, not all provinces’ Omicron onset timing fall within the uncertainty region of the fitted regression, suggesting that onset timing is only partially explained by within- or between-province colleague connections. To probe the association with case importation from international travel hubs, we therefore showed in supplementary Figure S5 that a ten-fold increase in connections with Noord-Holland was associated with a 7-day earlier on the onset timing of local outbreaks (95% CI: 0 to 14 days earlier), as Noord-Holland (NH) is estimated to experience the earliest Omicron likely due to international flights arriving at Amsterdam Airport Schiphol. Provinces with stronger colleague connections to Noord-Holland (NH), such as Zuid-Holland (ZH) and Utrecht (UT), tend to have earlier onsets, while more distant provinces like Zeeland (ZL) show later emergence.

### 2.3. Implications for regional interventions

The geographical granularity in the derived connectedness support evaluating potential impacts of regional interventions targeted at different administrative regions. A regional lockdown removes both colleague connections that originate from residents of the locked region and that occur in the shared workplace within that region. Figure 5 shows the effect on the spatial colleague connectivity per residential province pair (same structure as in Figure 2a) if different provinces are in lockdown. Each panel places one province under lockdown and maps the relative reduction of colleague connections per residential province pair. Panels are symmetric because colleague links are undirected, regardless of which residential province is listed first.

**Figure 5.**
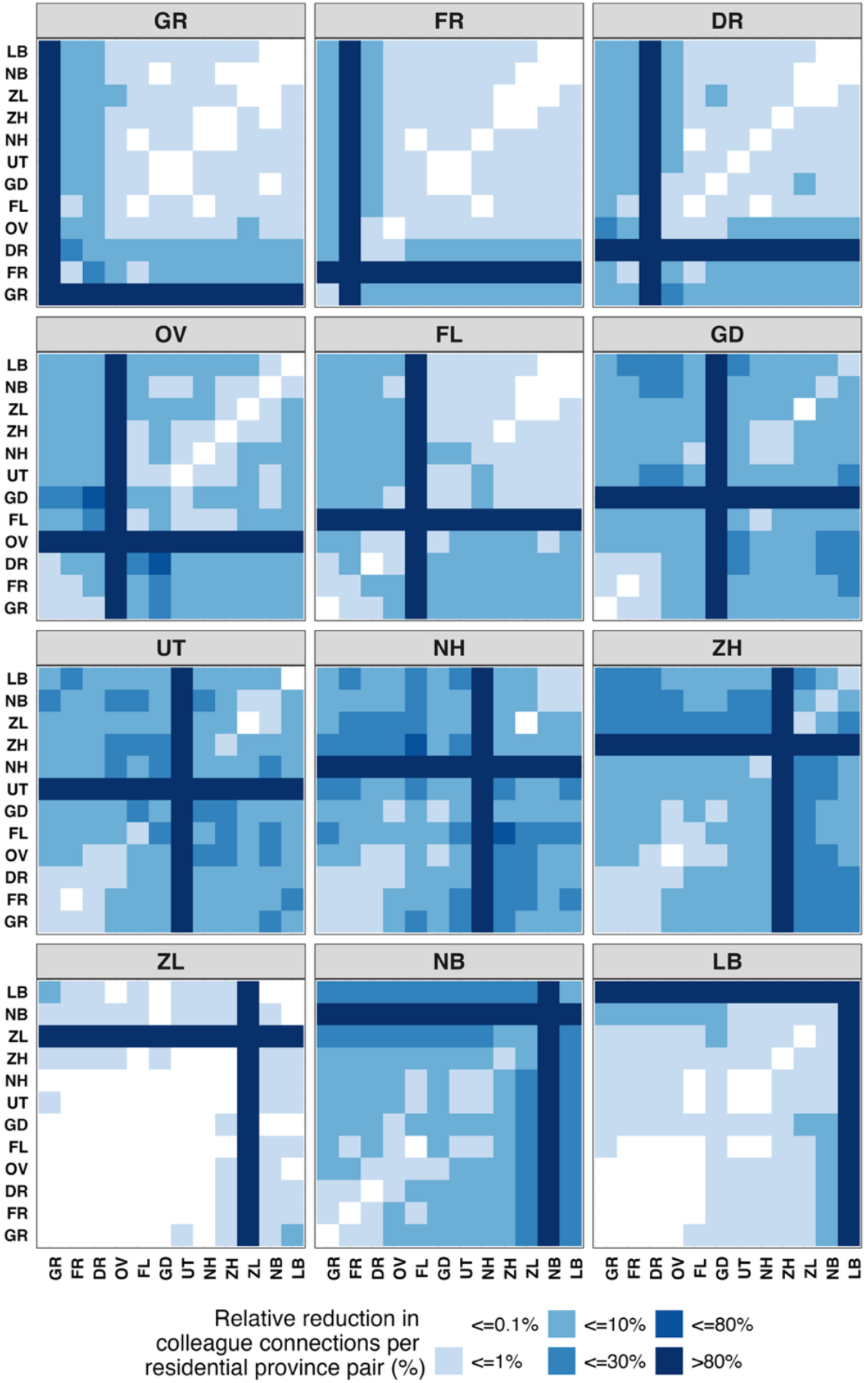
The potential impact of province-level lockdowns on colleague connectivity per residential province pair. Each panel visualizes the relative reduction in colleague connection between residential province pairs when the province labelled at the top of the panel is placed under lockdown. The darker the shades, the larger proportions of connections would be removed. Values were derived from Figure 3a.

As expected, colleague connections among residents of the locked province are entirely removed regardless of the meeting province, clearly shown by the darkest shades in all panels. Off-diagonal shading reflects connections between residents of two other provinces that meet inside the locked province. Lockdowns in Noord-Holland (NH), Zuid-Holland (ZH), and Utrecht (UT) affect the largest number of cross-province province pairs (dark off-diagonal cells), consistent with dense interprovincial connectivity centred on the Netherland’s western urban corridor (Randstad) around the country’s four largest cities. Notably, Gelderland (GD) and Noord-Brabant (NB) also exert national influence in connectedness patterns. The impact of simultaneous lockdowns in two or more provinces can be quantified using the same workflow. Analogous maps for safety-region lockdowns are provided in supplementary Figure S6 which demonstrate recognizably heterogenous impact of locking down one of the 25 Dutch safety regions.

After aggregating the removed connections to the national level (i.e. fractions of total colleague connections removed under a regional lockdown), the potential impacts of regional lockdowns remain uneven across targeted regions (Figure 6). Locking down one of the twelve provinces (blue bars) would remove only 2.6% (province ZL) to as much as 25.3% (province ZH) of all colleague connections. The range for safety regions (orange bars) is 2.5% to 12.5%. For municipalities (green bars), the potential impact is highly skewed. Locking down Amsterdam alone would remove 10.0% of all national colleague connections, while there are 305 municipalities each contributing less than 1% of all colleague connections. Municipalities with the top 15 contributions to colleague connections are labelled in green.

**Figure 6.**
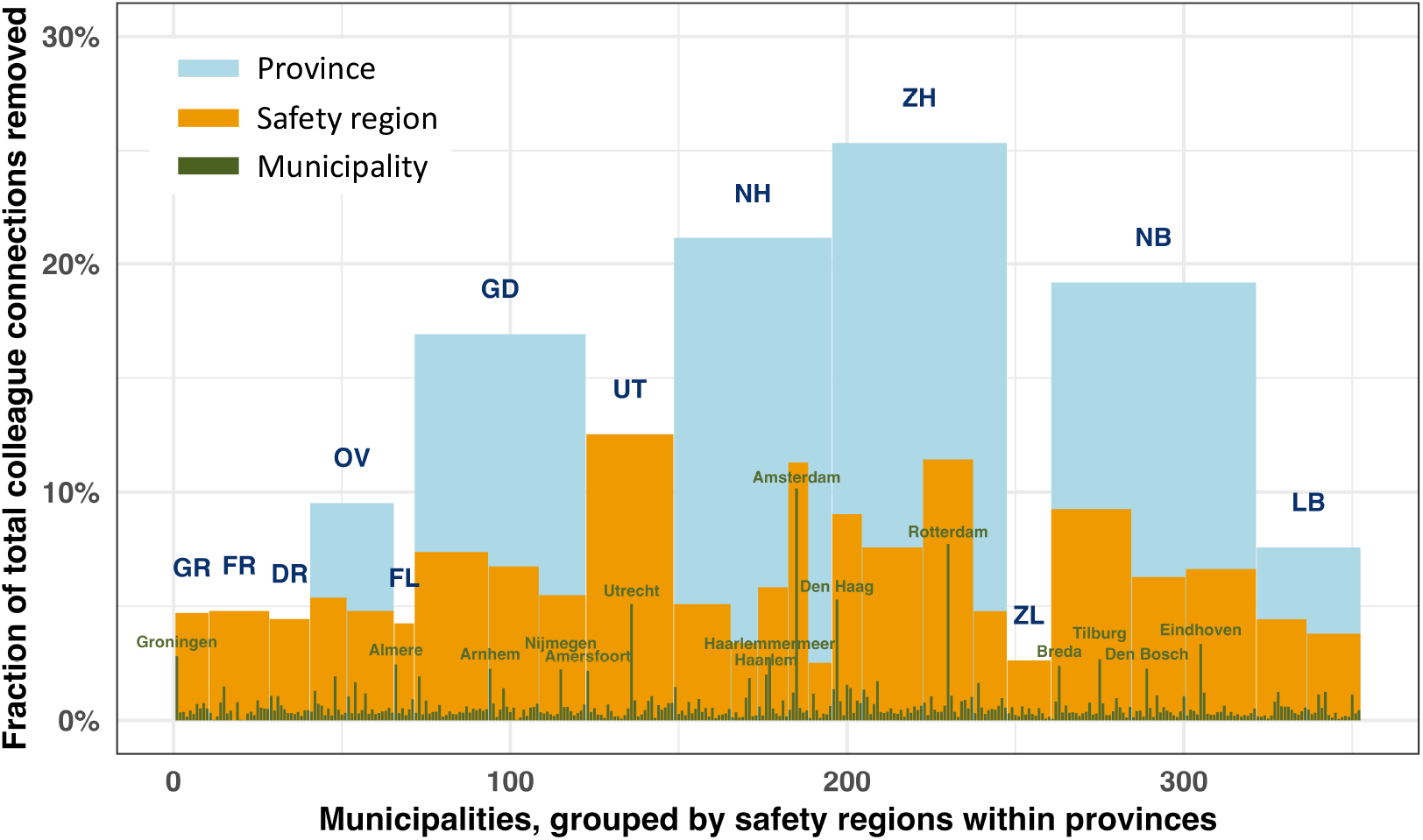
The potential impact of regional lockdowns on total national colleague connections. Each bar shows the share of all colleague connections that would be removed by locking down a single administrative unit at one of three levels: municipality (green), safety region (orange), or province (blue). The 352 municipalities are grouped by 25 safety regions within 12 provinces, using standard numbering from north to south. Province abbreviations are labelled in blue, while the municipalities with 15 largest contributions are labelled in green.

Though regional labour force size is one of the main drivers for a region’s contribution to national total colleague links, we successfully identified business hubs (where there are not necessarily many residents but a high concentration of companies) by including workplace municipality as a third spatial attribute. For instance, though municipality Haarlemmermeer has about one fourth less working population compared to municipality Almere (88.9 thousand vs. 119.1 thousand), locking down Haarlemmermeer would remove even more colleague connections than Almere (2.7% vs. 2.4%).

Cross-regional travel bans is an alternative regional intervention to the uniform regional lockdown, by removing only links that cross regional boundaries while preserving within-region colleague links. Under cross-region travel bans, the overall reductions in total colleague links (9.4% highest as shown in Figure 7) are consistently smaller compared to regional lockdowns (25.3% highest as shown in Figure 6). The contribution of labour force size is further downscaled when quantifying the reductions of total national colleague links. Supplementary Figure S7 shows that highly populated regions are not necessarily the most influential under cross-regional travel bans. For example, province Zuid-Holland (ZH) has about one fourth larger labour force than Noord-Holland (NH) (2.0 million vs. 1.6 million), but imposing a travel ban in ZH removes less connections than NH (9.1% vs. 9.4%), which is not the case for regional lockdowns (25.3% vs. 21.1%).

**Figure 7.**
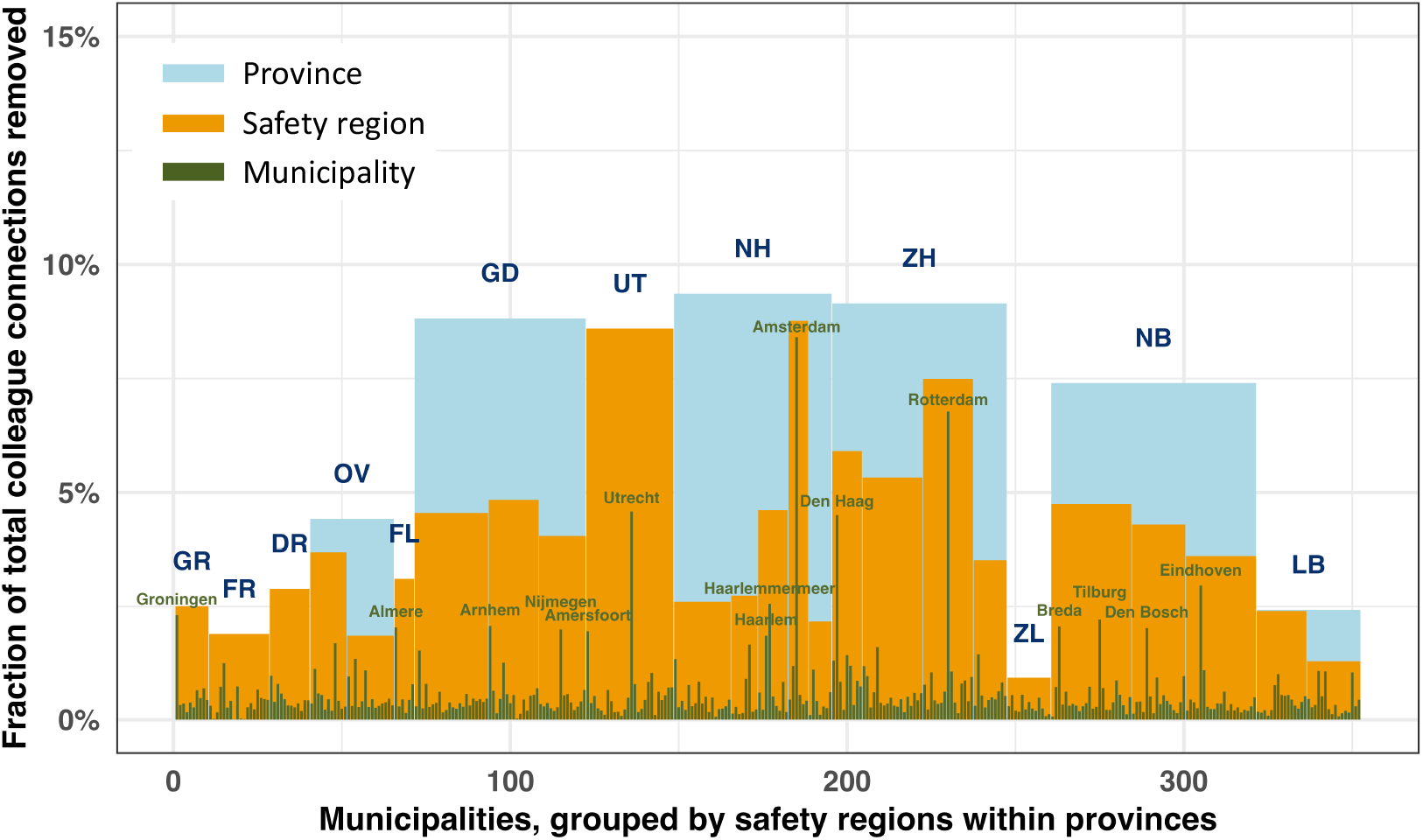
The potential impact of cross-regional travel bans on total national colleague connections. Each bar shows the share of all colleague connections that would be removed due to crossing the boundary of a single administrative unit (i.e., commuting into or out of that region). The rest of the captions are the same as Figure 6.

### 2.4. Sensitivity analysis

The spatial connectivity patterns are stable across different choices of the degree cap *D* (i.e. maximum number of colleagues an individual is allowed to have, shown in datasets presented online^23^). The ranking of provincial Omicron onset timing is stable across different choices of *e*_min_ (i.e. beyond which weekly incident cases defined the start of loglinear model, shown in supplementary Figure S4.1.) and to the threshold for onset *I* (i.e. beyond which weekly incident cases defined the emergence of Omicron, shown in supplementary Figure S4.2.).

## 3. Discussion

Our study presents a generalizable framework to translate individual-level registry data into geographical connectedness population contact networks, by pairing individuals working in the same local branch. We derived a highly fine-grained and broadly reusable data product which quantifies geographical connectedness of colleague networks within and between Dutch municipalities. We show that this connectedness can flag regions with elevated risks of a subsequent infection wave, by demonstrating associations between connectivity patterns and SARS-CoV-2 Omicron onset timing. We further showcased the utility of the derived data product in supporting regionally tailored interventions, using a regional lockdown and a cross-regional travel ban as examples.

Our geographical colleague connectedness corroborates the assumptions in recent studies^14,16^ that population mixing differs by geographic scale within a country. However, the share of region-specific mixing in these studies was assumed to be fixed and uniform across provinces, whereas our estimates show substantial interprovincial variations.

As for the provincial Omicron onset timings, Noord-Holland (NH) was estimated to experience the earliest Omicron onset regardless of different case thresholds. This clearly reflects elevated importation risk from international flights arriving at Amsterdam Airport Schiphol. Geographical colleague connectivity patterns explain part of the variation in provincial Omicron onset, but not all. For example, even though Flevoland (FL) is highly connected to Noord-Holland (NH), its Omicron onset was much later than the latter. This might be explained by the association analysis where Flevoland (FL) has relatively fewer within-province colleague connections. Another reason might be that, at the time of the Omicron outbreak, many companies in Noord-Holland (NH) had a working-from-home policy reducing the level of onsite mixing. In contrast, Gelderland (GD) shows a relatively early estimated Omicron onset, but it does not have that many colleague connections within-province, between-province, or to Noord-Holland (NH). A plausible explanation might be the substantial classmate connections around the university hub and cross-border mobility with Germany.

We are aware that part of the heterogeneous impact of a regional lockdown overlaps with the labour force size per administrative unit, as regions with more workers would have proportionally more working-age individuals that contribute largely to the national total colleague link counts. We also highlighted that the workplace locations in spatial connectedness enables identifying business-intensive regions by shifting leverage towards employment hubs. As such, our data product supports a better design and evaluation of subnational approaches compared to earlier work^18^, by allowing a regional intervention to target workplace locations without necessarily affect residence-areas.

We further argue that the proposed framework for translating microdata to spatial connectivity patterns can be generalized to other population contact settings or transferred beyond the Netherlands. The key prerequisite is access to individual-level registry data that include residential locations, linkages to relevant entities such as workplaces or schools and their geographic locations, and last but not least, legal and ethical access to the data. For example, apart from registrations on colleagues and companies, CBS also links students with their schoolmates, residents within the same households, and with their family members. There is ongoing work by our research team to characterize geographical connection patterns for student, family and household networks in the Netherlands, and investigate effectiveness of regional pandemic responses accordingly. For generalizability to international contexts, administrative data in the Nordic countries are systematically collected and have great potential to be linked, including microdata from Statistics Norway^24^, microdata from Statistics Denmark and microdata from Statistics Sweden^27^. With harmonized geographic units, standardized identifiers for individuals and entities, and our proposed framework, it is feasible to construct spatial connectivity for selected network layers in these countries at comparable municipal or aggregated geographical scales. Given that Norway and Sweden are 8 and 11 times larger in land area compared to the Netherlands, we expect stronger within-municipality social network connectivity, with fewer and more spatially fragmented long-distance connections. And a recent study from Denmark, where there is similar land area and cross-regional commuting tendencies, inferred plausible transmission clusters by linking registry social network data with sequencing data^28^, showing great potential of administrative data outside the Netherlands.

Our study has four limitations, for which we describe the consequences and planned mitigation steps below. First, in our two-stage Bayesian framework for estimating provincial Omicron onset, we used a plug-in approach by multiplying the posterior Omicron proportions by the total weekly COVID-19 cases and used this product as the rate parameter in the subsequent Poisson model. By doing so, only parameter uncertainty from the first stage is propagated to the second stage, while data uncertainty (i.e., the binomial sampling variability in Omicron cases) is omitted. This results in overconfident estimates of regional Omicron onset timing. Second, colleague connections in the context of this study do not imply real-world close contacts, due to lack of information on the frequency and duration of physical contacts per connection. It creates challenges when combining connections from different network layers as an average household contact is more likely to be more intimate and longer in period compared to a workplace contact. This gap could be partly addressed in future modelling work by incorporating insights on the relative weights of different types of social network connections from previous contact surveys. Third, current colleague connectivity has not explicitly included granularity on industrial sectors or by types of contracts, therefore limiting the design of possible intervention strategies (such as targeting on certain sectors). The level of granularity is partly constrained by CBS’s output requirements to minimize the risk of group disclosure. However, future work can focus on predefined subsets of sectors or contract types, thereby avoiding the need for fine-grained breakdowns across all sectors while still enabling more informative analyses within allowed disclosure limits. Fourth, it is important to note that the potential impact of regional interventions is based solely on proportion of (long-distance) colleague connections being removed. Future work is needed to first, integrate additional social network layers and second, quantify the epidemiological impact of regional interventions by incorporating these spatial connectivity patterns into transmission modelling frameworks.

In conclusion, we establish a reusable data product of geographical connectedness in colleague networks and confirm its utility in designing and evaluating more regionally targeted and social-network tailored pandemic measures. Also, we conclude that our workflow of inferring colleague networks and spatial connectivity patterns from individual-level registry data is valuable in its generalizability to other population contact settings and international contexts beyond the Netherlands.

## 4. Materials and methods

### 4.1. Overview of the methodology

This study integrates administrative data and epidemiological surveillance data to investigate the role of geographical connectivity in shaping subnational transmission dynamics during the Omicron wave in the Netherlands. Figure 8 provides an overview of the multi-step workflow and data sources which are discussed in the following sections. First (shown as Step A in Figure 8), we paired employees working for the same company by using employment relationships recorded in Dutch payroll tax registers and accessed via Statistics Netherlands (CBS) Microdata environment, following a similar methodology as Van der Laan et al. (2023)^29^. For large companies with local chain stores, such as supermarkets, people working at different branches are unlikely to have regular workplace contacts. Thus, we further refined this network by identifying employees of the same parent company who most likely work at the same local branch, using branch counts per branch location. Details of branch assignment algorithm can be found in Supplementary Information Algorithm S1. Then we linked individuals’ residential municipalities to the municipality where they work, to quantify the number of work-related connections per municipality triplet (two home municipalities for two individuals and their shared working municipality). The datasets with municipality-level geographical colleague connectivity are open access ^23^ together with the code for reproduction within CBS Microdata environment.

**Figure 8.**
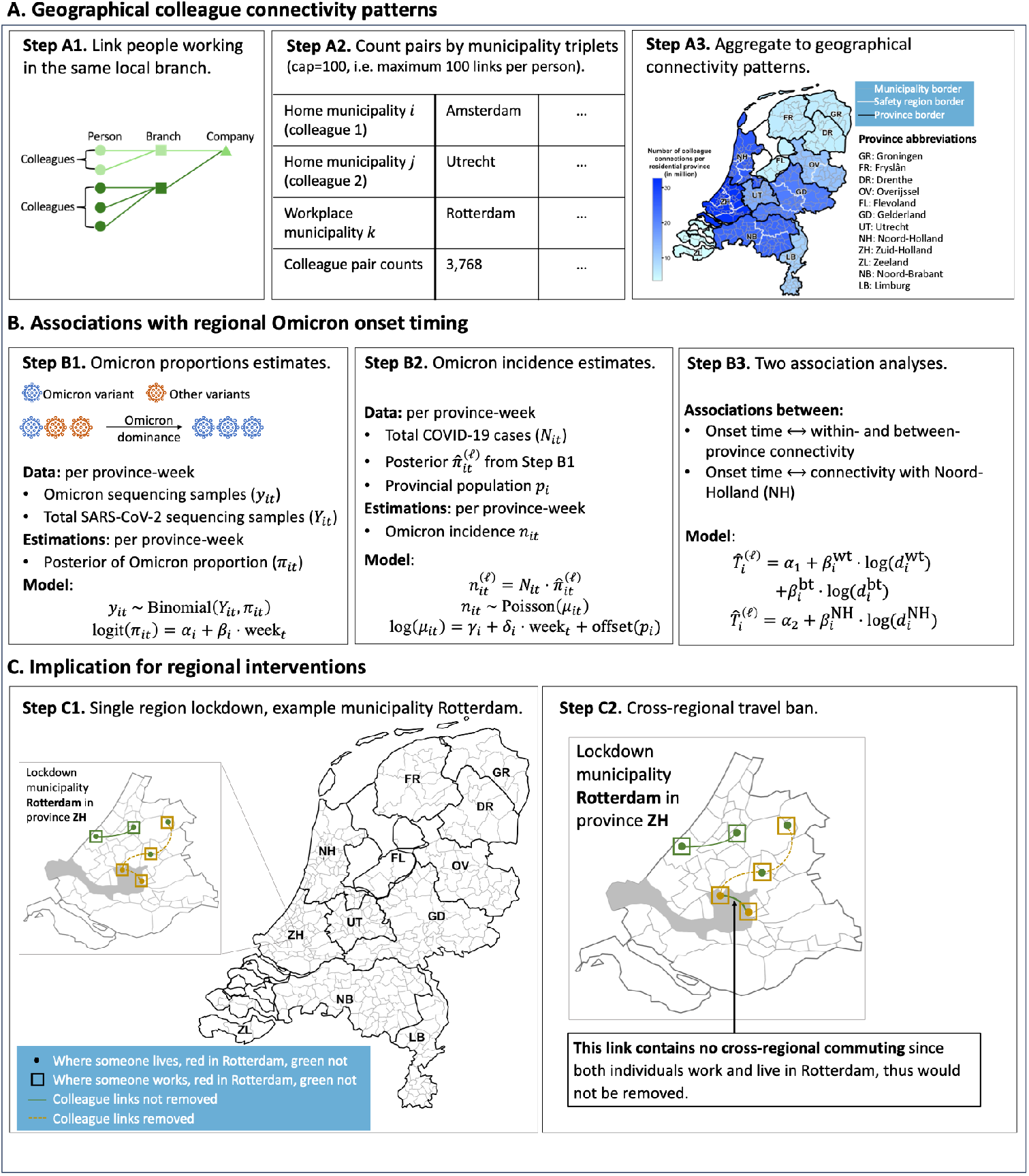
Overview of the data and methods. Our analysis framework consists of three steps: (A) construction of geographical colleague connectivity patterns across the Netherlands for its 352 municipalities, 25 safety regions, and 12 provinces; (B) estimation of provincial onset timing of SARS-CoV-2 Omicron variant, and evaluation of how this onset is associated with regional connectivity patterns; and (C) quantification of the impact of regional interventions on colleague connections within and between regions.

Second (shown as Step B in Figure 8), we estimated the onset timing of the SARS-CoV-2 Omicron variant per province by combining province-level case data from the National Institute for Public Health and the Environment |(RIVM) and variant-specific sequencing data from the Global Initiative on Sharing All Influenza Data (GISAID) ^30,31^. Then we quantified the associations between regional colleague connectivity and Omicron onset timing for each province. By doing so, we evaluated whether provinces with higher intra- or inter-province connectivity were associated with earlier onsets, and whether the time lag in onset relative to the first affected province was associated with the level of connectivity to that province.

Third and last (shown as Step C in Figure 8), we quantified the potential impact of regional interventions on total colleague links. Two regional interventions were implemented: (1) regional lockdowns, which would remove all colleague links that involve individuals living or working in the locked region; (2) cross-regional travel bans, which would remove all links that involve individuals commuting into or out of the targeted region.

### 4.2. Construction of geographical colleague connectivity patterns (Step A)

Here, we describe the algorithm and underlying assumptions to count the number of colleague links per municipality triplet 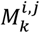, where *i* and *j* are the residential municipalities of two colleagues and *k* is their shared work municipality. These counts were then aggregated into metrics and matrices that characterize geographical patterns of colleague connectivity. First, individual-level administrative records were linked to group people employed by the same company. For companies with multiple branches within the same municipality, where data did not record which branch one employee works in, individuals were randomly allocated to branches to achieve approximately equal sizes. For companies with multiple branches within the same municipality, where the data did not specify the branch in which an employee worked, individuals were randomly assigned to branches while ensuring approximately equal branch sizes. Second, colleague links were counted for each branch and municipality triplet, aggregated across branches, and weighted by an assumed maximum number (cap) of colleagues per individual, with sensitivity analysis for this cap. Finally, links per municipality triplet were aggregated to summarize the spatial patterns in colleague connectivity across the Netherlands.

#### Registry data

This study uses person- and company-level registry data^32^ from Statistics Netherlands (CBS) covering the years 2019 to 2022, accessed via the secure CBS Microdata environment under approved data use agreements. Across the period of 2019 to 2022, these individual-level registry data cover 8.0 million (year 2019) to 8.5 million (year 2022) working individuals and 8.5 million (year 2019) to 9.0 million (year 2022) work income statements annually. Details of what these data entail and how these data were linked can be found in Supplementary Information Algorithm S1.

#### Link people working in the same branch (Step A1)

For companies that operate more than one branch within the same municipality, the registry data do not identify which local branch an individual is linked to. To avoid inflating colleague links by connecting individuals not co-located, we developed a branch-assignment algorithm to allocate each employee to the most probable branch and grouped colleagues accordingly, as follows. Let *N* be the number of employees working in the same municipality for a given company, and *B* the number of local branches in that municipality. Then the following three cases can be considered:

1. If *B* = 1, all *N* employees work at the same branch, so 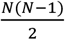 undirected colleague links in total.
2. If *B* > 1, *N* employees are deterministically and near-evenly allocated across *B* branches within the municipality. With near-even split, branch size *N*_*B*_ is either rounding down to the nearest integer 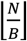 or rounding up to the nearest integer 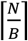, chosen such that the sizes sum exactly to the *N*. Within-branch colleague pairs can be computed accordingly (explained in equation 1).
3. If there is no information on *B*, we assume *B* = 1.

#### Count pairs by municipality triplet (Step A2)

After identifying co-workers working in the same local branch, individuals were clustered based on their home municipalities. Consider a branch *b* located in municipality *k* with *N*_*bk*_ employees. Let 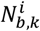 and 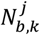 be the number of those employees who live in municipality *i* and *j*, respectively. Using undirected counting, unique colleague pairs within branch *b* for triplet (*i, j, k*) are represented as:

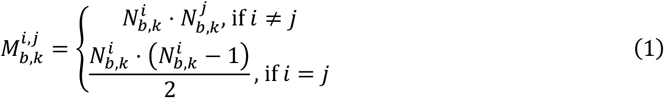

The symmetry 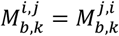 holds as pairs are undirected. Aggregating over all branches located in municipality *k* gives the total number of colleague pairs for triplet (*i, j, k*):

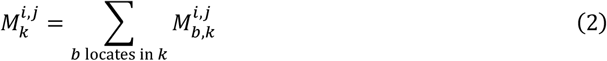

#### Degree cap

In large workplaces, individuals are expected to interact with only a subset of other employees working in the same branch office. We therefore let each employee have a degree cap *D* as the maximum number of colleague connections per individual is allowed to have. We followed the decision of Statistics Netherlands (CBS) and set as default: *D* = 100^29^. While this cap exceeds the average of 13.4 contacts per individual per day reported in diary-based contact studies ^5^, it accounts for potential cumulative contact opportunities over longer time periods.

Subsequently, for each branch *b* with *N*_*b*_ employees, we computed a weight based on the cap to scale the count of colleague pairs within branch *b* per municipality triplet:

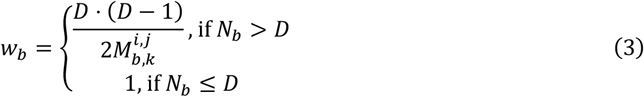

The total number of colleague pairs in equation (2) is weighted accordingly:

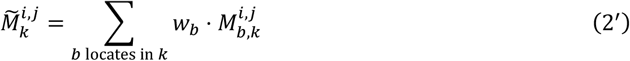

Supplement Algorithm S1 documents the details of how to assign colleagues to the most probable local branches and thereby count 131,910,029 colleague connections in total.

#### Aggregate to geographical connectivity patterns (Step A3)

In the Netherlands, municipalities, safety regions and provinces are the three regional administrative levels at which regional responses can be implemented^20,21^. We aggregated 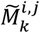, the number of colleague pairs per municipality triplet, to the safety region and province scales. The following summary statistics were derived to capture both the volume and share of colleague connections: (1) count of colleague connections by home-work or home-home province pairs; (2) a connection matrix by residential province pair with absolute connection counts, together with its row-normalized version to remove population-size scaling effects; (3) the share of connections that are within municipality, within province and between provinces.

### 4.3. Associations with regional Omicron onset timing (Step B)

In this section, we describe the data and framework to estimate the timing of Omicron emergence across Dutch provinces and relate this with geographical colleague connectivity patterns.

#### Sequencing and incidence data

We used two publicly available epidemiological datasets to estimate the timing of Omicron emergence per Dutch province. First, SARS-CoV-2 genome sequencing samples were downloaded from GISAID, a global data-sharing initiative for SARS-CoV-2 genomes and metadata^30^. These sequences were collected as part of the Dutch national germ surveillance program, in which Dutch laboratories mapped all the building blocks of the virus’ RNA to monitor variant prevalence and detect emerging variants of concern^33^. A total of 165,219 SARS-CoV-2 sequences collected from the 12 Dutch provinces were downloaded from GISAID, spanning from February 9^th^, 2020 to March 31^st^, 2023. By restricting the date starting from the week beginning December 28^th^, 2020, the first date when all 12 provinces had available genome sequences, until the week ending with March 31^st^, the end of sequence collection in the Netherlands, 157,891 sequences were left for analysis. To mitigate reporting fluctuations within a week, data were aggregated at the weekly level.

Second, municipality-level COVID-19 incidence was retrieved from the Dutch National Institute for Public Health and the Environment (RIVM) repository^33^, spanning from February 28, 2020, to March 31, 2023, the date when public testing facilities were closed. Data were aggregated to weekly new cases per province to avoid artificial fluctuations seen in daily case reporting and to align the time gap between case testing and sequencing.

#### Omicron proportions estimates (Step B1)

We then quantified provincial heterogeneity in the onset timing of Omicron emergence, defined as the first date when Omicron-positive weekly cases surpassed a pre-defined threshold. As an emerging variant after the Delta wave, Omicron exhibited substantial immune evasions which resulted in infections of individuals regardless of vaccination or infection with earlier variants^34,35^. The immune evasions of Omicron reduced bias due to varying immunity levels across provinces. Moreover, compared to earlier SARS-CoV-2 variants, Omicron emerged during a period when large-scale sequencing and testing infrastructure were in place in the Netherlands^33^. The higher surveillance capacity led to more complete and consistent data coverage across provinces, improving the credibility of Omicron onset timing estimates.

We developed a two-stage Bayesian inference workflow to simulate weekly Omicron cases over time. A hierarchical logistic regression model was first fitted to sequencing data. We modelled the number of Omicron-positive sequences *y*_*it*_ among all sequenced cases *Y*_*it*_ from province *i* in week *t* using a binomial likelihood,

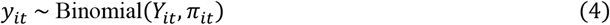

where *π*_*it*_ is the probability that a sequenced case was Omicron-positive. The log-odds of *π*_*it*_ were modeled with a linear predictor as:

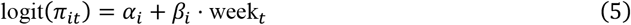

with *α*_*i*_ and *β*_*i*_ being province-specific intercepts and slopes, respectively. These parameters were given hierarchical priors, so that:

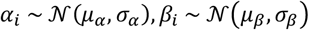

where hyperparameters (*μ*_*α*_, *μ*_*β*_, *σ*_*α*_, *σ*_*β*_) had vague priors specified in Supplementary Table S1. The model was fitted in Stan using Hamiltonian Monte Carlo (HMC) via the cmdstanr package (version 0.9.0). We ran 4 chains, each with 400 warm-up iterations followed by 400 sampling iterations.

#### Omicron incidence estimates (Step B2)

The weekly cases of Omicron *n*_*it*_ in province *i*, week *t*, was assumed to grow exponentially during the early phase, which is defined as the four-week period starting from the first week when weekly Omicron incidence exceeded a prespecified threshold *e*_min_, and continuing with an increase in the subsequent week’s incidence. The four-week window was chosen based on Grabowski et al. (2022)^36^.

Because *n*_*it*_ was unobserved, we imputed the expected values by multiplying the reported total COVID-19 new cases per week, *N*_*it*_, with the 1,600 posterior draws of the Omicron proportion 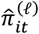 from the logistic model,

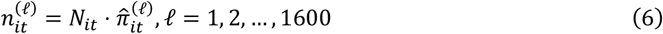

We then modelled the province-level weekly Omicron cases *n*_*it*_ using a Poisson generalized linear mixed model with a log link,

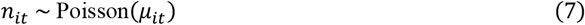

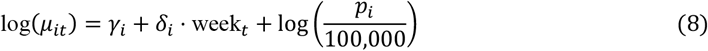

where *p*_*i*_ is provincial population size included as an offset, and province-specific intercepts *γ*_*i*_ and slopes *δ*_*i*_ were modeled hierarchically,

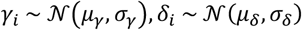

Each simulated series 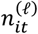 was then used as input to the model in equation (8), thereby propagating sequencing uncertainties to Omicron case estimates. We used a Poisson likelihood because the imputed 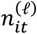 is a positive, continuous rate, and showed no evidence of overdispersion. This model was fitted using the brms package (version 2.22.0). We ran four Markov chains per imputed dataset, each with 1000 warm-up iterations followed by 1000 sampling iterations, thinned to keep every 4 iterations, resulting in 1.6 million total posterior draws.

#### Omicron onset timing

The onset timing of Omicron emergence per province was defined as the first week in which the predicted cases exceeded a fixed case threshold of *I* being 30. For province *i*, the onset time was derived for each posterior draw 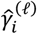 and 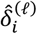 as:

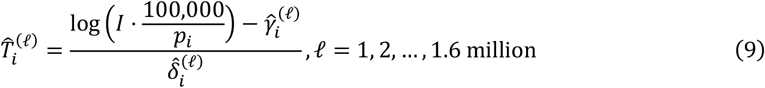

#### Two associations analyses (Step B3)

We did a Bayesian regression analysis to validate to which extent regional Omicron onset timing is associated with the geographical colleague connectivity patterns. From the full posterior of 1.6 million draws of onset times 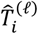, we randomly sampled 1000 draws and fit a Bayesian regression to each of the sample as indicated below.

For each province *i*, we summarized the connectivity derived in Section 2.2 to quantify the level of internal and external links. We decomposed links per residential province into two directional categories: (1) within-province links where both individuals live and work in province *i*, denoted as 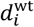; (2) between-province links where at least one individual resides or works in another province, denoted as 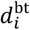. Because multiplicative changes are more meaningful compared to additive changes in colleague connections, we worked with a logarithmic scale of the two types of connections and then standardized across provinces for better Hamilton Monte Carlo sampling efficiency,

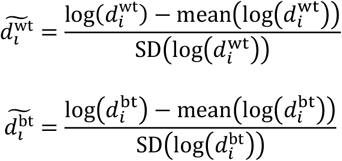

We then fit the model for each draw of the estimated Omicron onset time for province *i*

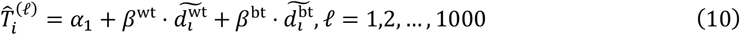

As Omicron was first seeded in the Netherlands through international flights from South Africa to the Netherlands, the onset of Omicron in Noord-Holland, where Amsterdam Airport Schiphol locates, was driven by international transmissions instead of within-country mixing. We additionally tested whether stronger links to Noord-Holland were associated with earlier local onset elsewhere. We computed for each residential province, number of colleague connections with individuals living in Noord-Holland, denoted as 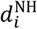. Similarly, we first standardized the logarithmic of 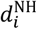 for sampling efficiency:

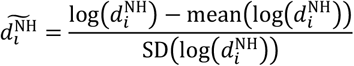

We fit the second model as:

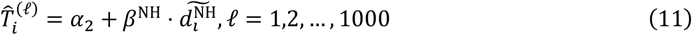

### 4.4. Implications for regional interventions (Step C)

Finally, we investigated how the geographical colleague connectivity patterns would change if two different regional interventions were in place. This potential impact of targeted interventions was quantified by the absolute count and proportion of colleague connections removed, both nationally and by residential province pair. If a region is in uniform lockdown, we removed all colleague links where at least one individual lived or worked in the targeted region. Alternatively, a cross-regional travel ban would only remove links involving individuals commuting into, or out of the targeted region, while keeping the links where both home and workplace were within that region. This travel-ban therefore preserves within-region workplace connectivity, which may allow essential onsite activities such as healthcare work and local supermarket supply to continue.

To assess how regional workforce size influences the potential intervention impact, we regressed the fraction of removed colleague links on regional workforce size. Regional workforce sizes were calculated by regional population aged over 15 years minus the unemployment counts, both accessed via CBS Open Data portal^37^.

### 4.5. Sensitivity analyses

The cap *D* (i.e. the maximum number of colleagues an individual is allowed to have) were chosen from the values of 50, 100 and 200 for year 2019 and 2022. The threshold *e*_min_, above which the start of the 4-week period to fit a loglinear increase in incident cases was defined, were chosen from 1, 3, 5 and 10. The case threshold *I*, above which the onset timing of Omicron emergence was defined, were chosen from 10, 20, 30, 100, 200 and 300.

### 4.6. Ethics statement

This study used registry data on individuals and companies from the Statistics Netherlands (CBS). Formal ethical approval for the use and publication of these data in the context of the PDPC-SURE Frontrunner project was obtained from the Erasmus MC Medical Ethical Council, under registration number MEC-2025-0338. Additionally, the research question and the required datasets were reviewed by CBS for feasibility and compliance with the General Data Protection Regulation (GDPR). Data access was provided exclusively through the secure CBS Microdata environment. All outputs from registry data were checked by CBS through a disclosure risk assessment before being released from the secure environment.

## Supporting information

Supplementary material with tables and figures.

## Acknowledgements

This work was funded by Pandemic and Disaster Preparedness Center (PDPC), grant number 2022.004. We also gratefully acknowledge all data contributors, i.e. the authors, the originating and submitting laboratories for the genome sequences in GISAID’s EpiCoV database. We thank CBS Microdata team for their contributions on collection, management, and expert consultations on the registry data of the working population and companies in the Netherlands. PPS is further grateful for Prof. Dr. Frank P. Pijpers for his critical information on availability of registry data in other European countries, and to Dr. Harrison Quick for extensive and constructive discussions of the Bayesian statistical framework.

## Code and data availability

The individual-level administrative data are not publicly available and can only be accessible upon request to CBS Microdata. The aggregated colleague connectivity (i.e. colleague pair counts per municipality triplet that this study generates) is openly accessible, together with the code that could reproduce the data within CBS Microdata environment^23^. Genome sequences and associated metadata are published in GISAID’s EpiCoV database with the GISAID identifier being EPI_SET_251126rq, and openly accessible^38^. Population size over 15 years old and population unemployed per Dutch municipality were accessed via CBS OpenData portal^37^. R code for Omicron onset estimations, association analyses and regional intervention scenarios is available at https://gitlab.com/erasmusmc-public-health/colleaguenet.

## Author contributions

Conceptualization: All authors.

Data analysis: PingPing Song.

Methodology - population connectivity: All authors.

Methodology - epidemiology: PingPing Song, Sake J. de Vlas, Luc E. Coffeng.

Visualization: PingPing Song, Sake J. de Vlas, Luc E. Coffeng.

Writing - original draft: PingPing Song.

Writing - review & editing: All authors.

