## Supplementary material with tables and figures. for "Mapping spatial colleague connectivity patterns from individual-level registry data to inform regional pandemic interventions"

**Contents**

**Algorithm S1.** Quantify geographical colleague connectivity by linking registry data files via CBS Microdata environment and assigning colleagues to local branches.

**Table S1.1.** Number of colleague connections per residential province pair (logarithmic scale with 10 base, raw data used in Figure 2a).

**Table S1.2.** Number of total colleague connections per residential province (original and logarithmic scale with 10 base, raw data used in Figure 2b).

**Table S1.3.** Row-normalized colleague connections per residential province (raw data used in Figure 2c).

**Table S2.** Colleague connectivity shares by province and geographic scale (raw data used in Figure 3).

**Table S3.** Priors used in Bayesian analysis.

**Figure S1.** Probability of Omicron in sequenced SARS-CoV-2 cases per province.

**Figure S2.** Weekly Omicron-positive cases per province ($e_{\min} = 1$).

**Figure S3.** Estimated Omicron onset time per province (with the case threshold $I = 30$).

**Figure S4.1.** Sensitivity of Omicron onset time to the start of loglinear regression $e_{\min}$.

**Figure S4.2.** Sensitivity of Omicron onset time ranking to the case threshold $I$.

**Figure S5.** Association between connectivity with Noord-Holland and Omicron onset timing.

**Figure S6.** Potential impact of safety-region-level lockdowns on colleague connections per residential safety-region pair.

**Figure S7.** Comparison of the potential impact of two regional interventions on total national colleague connections.

**Algorithm S1.** Quantify geographical colleague connectivity by linking registry data files via CBS Microdata environment and assigning colleagues to local branches.

**Step 1**. Load registry data files from Statistics Netherlands (CBS) Microdata.

Three types of data files are loaded within CBS Microdata environment, they are: (1) job relationships (i.e., **SPOLISBUS**), (2) individuals' home addresses (i.e., **GBAADRESOBJECT**, **VSLGTAB** and **GIN**), and (3) information on local branches (i.e., **GEMEENTESTPLTAB** and **ABR_REGIO**).

- **SPOLISBUS**: This is a database containing information on jobs and wages of employees at Dutch companies, available from the year 2010 onward. Each row in the database represents an income relationship, uniquely identified by the variable *IKVID*. Alongside *SBEID* (a unique identifier for an employer) and *RINPERSOON* (a unique identifier for an individual), each row captures the employment connection between an employer and an individual.
- **GBAADRESOBJECT**: This database contains the addresses of individuals registered with the municipality in the Personal Records Database. Each row links an individual (identified by the variable *RINPERSOON*) to a residential object (identified by a unique, meaningless number, *RINOBJECTNUMMER*). This residential object can be further linked to *VSLGTAB* to obtain the actual addresses. These data are available from year 1995 onwards upon request to CBS.
- **VSLGTAB**: This database contains the municipality code for inhabit anted objects, linked to their unique identifiers *RINOBJECTNUMMER*. Data are available from 1995 onwards.
- **GIN (GebiedeninNederland)**: This data file shows relationship between Dutch municipal divisions with higher-level administrative divisions of the Dutch territory. It merges municipal divisions with safety regions, which are administrative areas established to coordinate and manage emergency preparedness, response, and public safety measures.
- **GEMEENTESTPLTAB**: This database contains the municipality code associated with each job as of December in the reporting year, available from 2014 onward. Similar to SPOLISBUS, each row is uniquely identified by an income relationship (*IKVID*), along with identifiers for the person (*RINPERSOONS*) and their employer (*SBEID*). If multiple local branches belong to the same head company, only one *SBEID* is assigned. This database enhances the algorithm by providing additional information on an individual's working municipality (*GEMEENTESTPL*). Consequently, the algorithm can establish connections among individuals working in the same local branch, rather than grouping them solely under the same head company.

- **ABR_REGIO**: This database provides additional, fine-grained information on companies. Specifically, it includes the number of local branches within the same municipality, recorded in the variable *Vest_Gemeente*.

**Step 2.** Merge loaded data files.

The loaded data files were linked using unique identifiers that are consistent across the databases. Through this linking process, information from multiple databases is combined and organized into a single data frame for further analysis.

- Step 2.1. **SPOLISBUS** was merged with **GBAADRESOBJECT** by linking the variable *RINPERSOON*, resulting in **SPO_GBA**. In this merged dataset, each individual in the colleague database is identified by a residential object number (i.e., *RINOBJECTNUMMER*).
- Step 2.2. **SPO_GBA** was merged further with **VSLGTAB** and **GIN** by linking *RINOBJECTNUMMER* and municipality code respectively, resulting in **SPO_ADR** where each individual in colleague database has information on home municipality and safety region.
- Step 2.3. To gather more detailed information on company structures, especially on which local branch each individual is (most probably) working in, **SPOLISBUS** was merged with **GEMMENTESTPLTAB** and **ABR_REGIO**. Company-level information is linked through mapping the unique job identification (i.e., *IKVID*). This merging process resulted in **SPO_ADR_LBE**, where each row represents a job relationship along with information on the individual's home address (at both the municipality and safety region levels), working municipality, and the number of local branches belonging to the same head company within the same municipality.

**Step 3.** Pseudo code to compute colleague links per municipality triplet by assigning colleagues to local branches.

---

**Input:**

$(i, j, k)$: home municipalities $i, j$ and work municipality $k$

$\{C_k\}$: set of head companies with at least one employee working in $k$

$\{B_k^C\}$: set of local branches of company $C_k$ and operate in $k$

$B_k^C = |\{B_k^C\}|$: number of branches of company $C$ and operate in $k$

$N_k^C$: total number of employees of $C$ and work in $k$ (i.e., $N_k^C \geq 1$)

$D$: maximum number of colleague links per individual is allowed to have (default $D = 100$)

**Output:**

$M_k^{i,j}$: number of colleague pairs where colleagues live in municipalities $i$ and $j$, and work in $k$

**for** a given municipality triplet $(i, j, k)$:

  **for** each company $C_k \in \{C_k\}$

    **if** $\left(N_k^C == 1\right)$

      next

    **end if**

    **if** (is.na$\left(B_k^C\right)$){

      $B_k^C \leftarrow 1$}

    **end if**

    Assign each of the $N_k^C$ employees a branch label $b \in \{1, \dots, B_k^C\}$ using random permutation of the vector formed by repeating $1, \dots, B_k^C$ to length $N_k^C$

    **for** $\left(b = 1, \dots, B_k^C\right)${

      $n_{b,k} \leftarrow$ number of employees assigned to branch $b$

      Count employees living in municipality $i$ or $j$ as $n_{b,k}^i$ or $n_{b,k}^j$

      Compute colleague pairs by municipality triplet as:

$$M_{b,k}^{i,j} = \begin{cases} n_{b,k}^i \cdot n_{b,k}^j, \text{ if } i \neq j \\ \dfrac{n_{b,k}^i \cdot \left(n_{b,k}^i - 1\right)}{2}, \text{ if } i = j \end{cases}$$

      Compute branch weight based on degree cap $D$:

$$w_b = \begin{cases} \dfrac{D \cdot (D-1)}{2M_{b,k}^{i,j}}, \text{ if } n_{b,k} > D \\ 1, \text{ if } n_{b,k} \leq D \end{cases}$$

    **end for**

  **end for**

  Aggregate over all branches $b$ with its head company $C_k \in \{C_k\}$

$$\widetilde{M}_k^{i,j} = \sum_b w_b \cdot M_{b,k}^{i,j}$$

**end for**

---

**Table S1.1. Number of colleague connections per residential province pair (logarithmic scale with 10 base, raw data used in Figure 2a).** This table presents the raw data used for generating Figure 2a, showing the number of colleague connections between residential province pairs. The data is transformed to a logarithmic scale (log base 10) to better visualize the heterogeneity between provinces. The order of the provinces in rows and columns are the same as how it is presented in Figure 2a. Full names of provinces and the corresponding abbreviations are shown in supplementary Table S1.2.

| | **GR** | **FR** | **DR** | **OV** | **FL** | **GD** | **UT** | **NH** | **ZH** | **ZL** | **NB** | **LB** |
|---|---|---|---|---|---|---|---|---|---|---|---|---|
| **LB** | 4.39 | 4.46 | 4.44 | 4.88 | 4.48 | 5.65 | 5.11 | 5.26 | 5.38 | 4.45 | 6.20 | 6.84 |
| **NB** | 4.84 | 4.95 | 4.89 | 5.34 | 5.04 | 6.29 | 5.87 | 5.83 | 6.35 | 5.67 | 7.20 | 6.20 |
| **ZL** | 3.99 | 4.11 | 3.98 | 4.31 | 4.05 | 4.71 | 4.59 | 4.79 | 5.59 | 6.36 | 5.67 | 4.45 |
| **ZH** | 5.08 | 5.14 | 5.01 | 5.41 | 5.46 | 5.96 | 6.30 | 6.53 | 7.34 | 5.59 | 6.35 | 5.38 |
| **NH** | 5.19 | 5.40 | 5.03 | 5.46 | 6.18 | 5.91 | 6.40 | 7.20 | 6.53 | 4.79 | 5.83 | 5.26 |
| **UT** | 4.90 | 4.92 | 4.86 | 5.46 | 5.67 | 6.39 | 6.75 | 6.40 | 6.30 | 4.59 | 5.87 | 5.11 |
| **GD** | 5.04 | 5.13 | 5.25 | 6.32 | 5.68 | 7.06 | 6.39 | 5.91 | 5.96 | 4.71 | 6.29 | 5.65 |
| **FL** | 4.62 | 5.20 | 4.82 | 5.58 | 6.22 | 5.68 | 5.67 | 6.18 | 5.46 | 4.05 | 5.04 | 4.48 |
| **OV** | 5.21 | 5.43 | 5.89 | 6.85 | 5.58 | 6.32 | 5.46 | 5.46 | 5.41 | 4.31 | 5.34 | 4.88 |
| **DR** | 6.13 | 5.59 | 6.35 | 5.89 | 4.82 | 5.25 | 4.86 | 5.03 | 5.01 | 3.98 | 4.89 | 4.44 |
| **FR** | 5.81 | 6.60 | 5.59 | 5.43 | 5.20 | 5.13 | 4.92 | 5.40 | 5.14 | 4.11 | 4.95 | 4.46 |
| **GR** | 6.49 | 5.81 | 6.13 | 5.21 | 4.62 | 5.04 | 4.90 | 5.19 | 5.08 | 3.99 | 4.84 | 4.39 |

**Table S1.2. Number of total colleague connections per residential province (original and logarithmic scale with 10 base, raw data used in Figure 2b).** This table presents the raw data used for generating Figure 2b, both the original and logarithmic scale with a base of 10.

| Province | Number of all colleague connections *(in million)* | Logarithmic scale with base 10 |
|---|---|---|
| **Groningen (GR)** | 5.86 | 6.77 |
| **Fryslân (FR)** | 6.14 | 6.79 |
| **Drenthe (DR)** | 5.41 | 6.73 |
| **Overijssel (OV)** | 11.88 | 7.07 |
| **Flevoland (FL)** | 5.21 | 6.72 |
| **Gelderland (GD)** | 21.03 | 7.32 |
| **Utrecht (UT)** | 14.46 | 7.16 |
| **Noord-Holland (NH)** | 25.90 | 7.41 |
| **Zuid-Holland (ZH)** | 32.12 | 7.51 |
| **Zeeland (ZL)** | 3.41 | 6.53 |
| **Noord-Brabant (NB)** | 24.23 | 7.38 |
| **Limburg (LB)** | 9.76 | 6.99 |

**Table S1.3. Row-normalized colleague connections per residential province (raw data used in Figure 2c).** This table presents the raw data used for generating Figure 2c, showing the relative distribution of colleague connections from each residential province to another province. The data is transformed to a logarithmic scale with base 10 to better visualize the heterogeneity between provinces. The order of the provinces in rows and columns are the same as how it is presented in Figure 2c. Full names of provinces and the corresponding abbreviations are shown in supplementary Table S1.2.

| | **GR** | **FR** | **DR** | **OV** | **FL** | **GD** | **UT** | **NH** | **ZH** | **ZL** | **NB** | **LB** |
|---|---|---|---|---|---|---|---|---|---|---|---|---|
| **LB** | 0.003 | 0.003 | 0.003 | 0.008 | 0.003 | 0.046 | 0.013 | 0.019 | 0.025 | 0.003 | 0.162 | 0.713 |
| **NB** | 0.003 | 0.004 | 0.003 | 0.009 | 0.005 | 0.081 | 0.031 | 0.028 | 0.092 | 0.019 | 0.661 | 0.065 |
| **ZL** | 0.003 | 0.004 | 0.003 | 0.006 | 0.003 | 0.015 | 0.011 | 0.018 | 0.115 | 0.676 | 0.138 | 0.008 |
| **ZH** | 0.004 | 0.004 | 0.003 | 0.008 | 0.009 | 0.028 | 0.062 | 0.105 | 0.688 | 0.012 | 0.069 | 0.007 |
| **NH** | 0.006 | 0.010 | 0.004 | 0.011 | 0.058 | 0.031 | 0.096 | 0.618 | 0.131 | 0.002 | 0.026 | 0.007 |
| **UT** | 0.005 | 0.006 | 0.005 | 0.020 | 0.032 | 0.171 | 0.388 | 0.172 | 0.137 | 0.003 | 0.052 | 0.009 |
| **GD** | 0.005 | 0.006 | 0.008 | 0.099 | 0.023 | 0.542 | 0.117 | 0.038 | 0.043 | 0.002 | 0.093 | 0.021 |
| **FL** | 0.008 | 0.031 | 0.013 | 0.072 | 0.321 | 0.093 | 0.090 | 0.288 | 0.056 | 0.002 | 0.021 | 0.006 |
| **OV** | 0.014 | 0.023 | 0.066 | 0.594 | 0.032 | 0.176 | 0.024 | 0.024 | 0.022 | 0.002 | 0.018 | 0.006 |
| **DR** | 0.248 | 0.072 | 0.419 | 0.144 | 0.012 | 0.033 | 0.013 | 0.020 | 0.019 | 0.002 | 0.014 | 0.005 |
| **FR** | 0.104 | 0.642 | 0.063 | 0.044 | 0.026 | 0.022 | 0.014 | 0.041 | 0.022 | 0.002 | 0.015 | 0.005 |
| **GR** | 0.531 | 0.109 | 0.228 | 0.028 | 0.007 | 0.019 | 0.013 | 0.026 | 0.020 | 0.002 | 0.012 | 0.004 |

**Table S2. Colleague connectivity shares by province and geographic scale (raw data used in Figure 3).** This table presents the raw data used to plot Figure 3 in the paper, which shows the absolute numbers and proportions of colleague connections originating from individuals living within the same municipality (i.e. within municipality), different municipalities within the same province (i.e. within province), and different provinces (i.e. between provinces). Full names of provinces and the corresponding abbreviations are shown in supplementary Table S1.2.

| **Province** | **n(all connections)** *in million* | **n(within municipality)** *in million* | **n(within province)** *in million* | **n(between provinces)** *in million* | **%(within municipality)** | **%(within province)** | **%(between provinces)** |
|---|---|---|---|---|---|---|---|
| **GR** | 5.86 | 1.51 | 1.60 | 2.75 | 25.7% | 27.4% | 46.9% |
| **FR** | 6.14 | 1.64 | 2.30 | 2.20 | 26.7% | 37.6% | 35.8% |
| **DR** | 5.41 | 1.16 | 1.10 | 3.14 | 21.5% | 20.4% | 58.2% |
| **OV** | 11.88 | 2.98 | 4.08 | 4.83 | 25.1% | 34.3% | 40.6% |
| **FL** | 5.21 | 1.23 | 0.45 | 3.54 | 23.6% | 8.6% | 67.9% |
| **GD** | 21.03 | 4.20 | 7.20 | 9.64 | 20.0% | 34.2% | 45.8% |
| **UT** | 14.46 | 2.35 | 3.27 | 8.84 | 16.2% | 22.6% | 61.2% |
| **NH** | 25.90 | 6.05 | 9.94 | 9.90 | 23.4% | 38.4% | 38.2% |
| **ZH** | 32.12 | 7.23 | 14.86 | 10.03 | 22.5% | 46.3% | 31.2% |
| **ZL** | 3.41 | 0.98 | 1.33 | 1.10 | 28.8% | 38.9% | 32.4% |
| **NB** | 24.23 | 5.35 | 10.65 | 8.22 | 22.1% | 44.0% | 33.9% |
| **LB** | 9.76 | 2.18 | 4.78 | 2.80 | 22.3% | 49.0% | 28.7% |

**Table S3. Priors used in Bayesian analysis.** Priors used for Bayesian inference framework to estimate provincial Omicron onset timing (i.e. logistic regression model and loglinear regression model), and the association analyses between Omicron onset and geographical colleague connectivity patterns.

| | | |
|---|---|---|
| **Logistic regression model** | | |
| $\mu_\alpha$ | Mean of provincial intercepts | Normal (1, 2) |
| $\sigma_\alpha$ | Standard deviation of provincial intercepts | Exponential (1) |
| $\mu_\beta$ | Mean of provincial slopes | Normal (5, 2) |
| $\sigma_\beta$ | Standard deviation of provincial slopes | Exponential (1) |
| **Loglinear regression model** | | |
| $\mu_\gamma$ | Mean of provincial intercepts | Normal (0, 0.1) |
| $\sigma_\gamma$ | Standard deviation of provincial intercepts | Exponential (1) |
| $\mu_\delta$ | Mean of provincial slopes | Normal (1, 0.5) |
| $\sigma_\delta$ | Standard deviation of provincial slopes | Exponential (1) |
| **Association analysis of within- and between-province connections** | | |
| $\alpha_1$ | Intercept (week number) | Normal (46, 2) |
| $\beta^{\mathrm{wt}}$ | Effect of standardized log within-province connections | Normal (−0.5, 0.5) |
| $\beta^{bt}$ | Effect of standardized log between-province connections | Normal (−0.5, 0.5) |
| $\sigma_1$ | Standard deviation of the residuals | Exponential (2) |
| **Association analysis of connections with Noord-Holland (NH)** | | |
| $\alpha_2$ | Intercept (difference in weeks) | Normal (3, 0.5) |
| $\beta^{NH}$ | Effect of standardized log connections to Noord-Holland (NH) | Normal (−0.5, 0.5) |
| $\sigma_2$ | Standard deviation of the residuals | Exponential (2) |

**Figure S1. Probability of Omicron in sequenced SARS-CoV-2 cases per province.** Observed proportions (green circles) represent the number of Omicron-positive sequences divided by the total number of sequenced SARS-CoV-2 cases per province. Posterior means and 95%CI from logistic regression model are shown as red lines and ribbons.

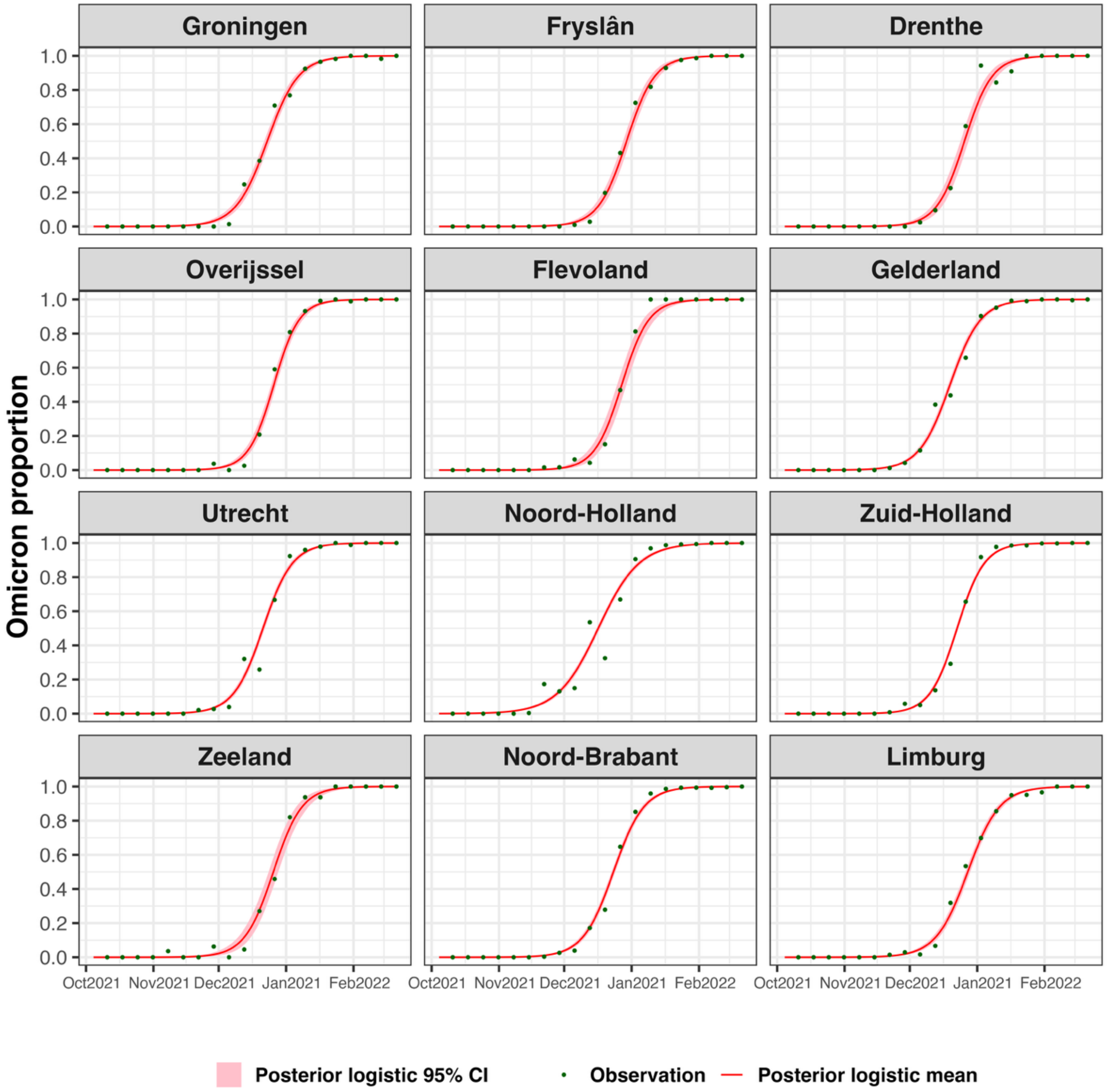

**Figure S2. Weekly Omicron-positive cases per province ($e_{\min} = 1$).** The estimated Omicron-positive incidence across provinces and time. The circles (red) and lines (blue) represent the posterior averages, the error bars (red) and ribbon areas (blue) show the 95% credible intervals for the logistic and loglinear model respectively. The start of the loglinear regression per province is defined as the first week when the posterior mean of expected Omicron new cases exceeded $e_{\min} = 1$. Lower bounds of the red bars were added by 0.1 if being 0, to allow meaningful logarithmic scale in the y axis.

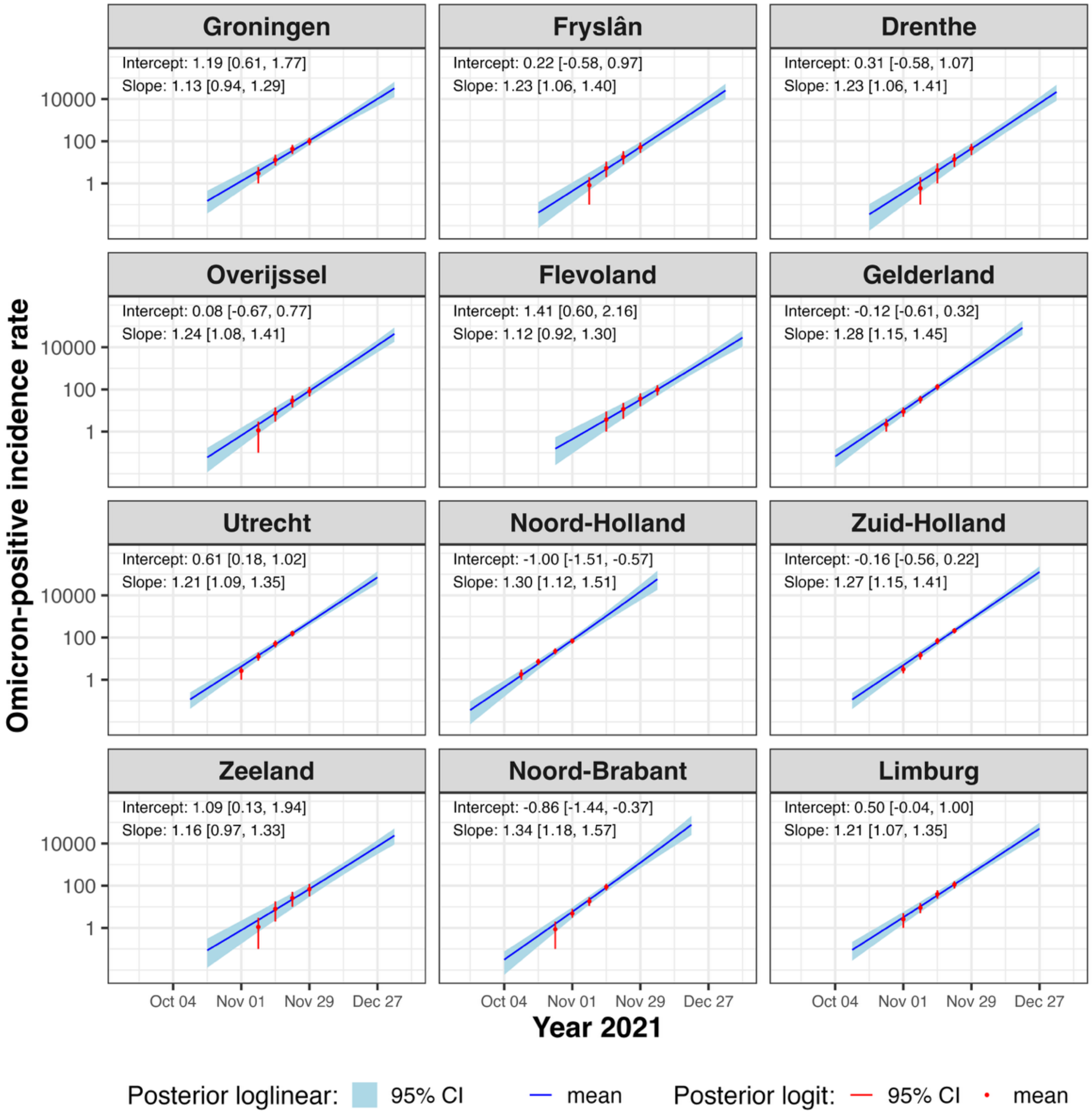

**Figure S3. Estimated Omicron onset time per province (with the case threshold $I = 30$).** Posterior means (circles) and 95% credible intervals (error bars) indicate the estimated date in year 2021 when each province first reached 30 Omicron-positive cases.

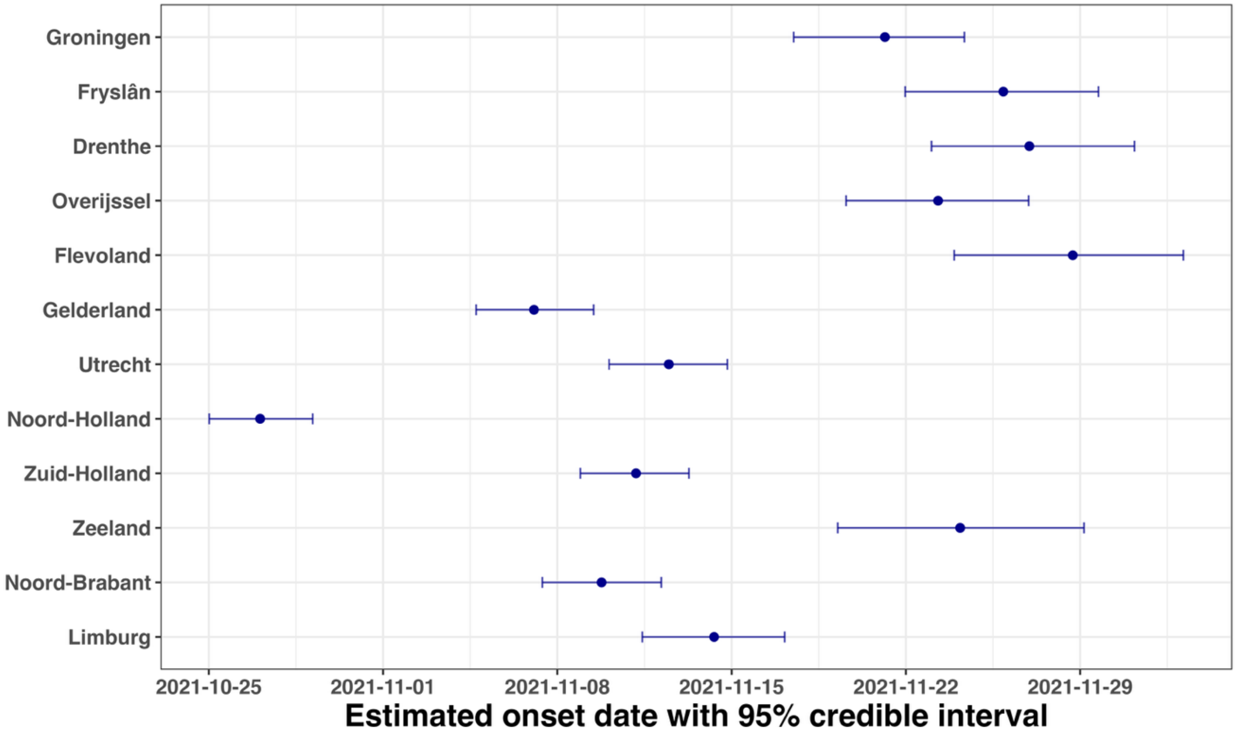

**Figure S4.1. Sensitivity of Omicron onset time to the start of loglinear regression $e_{min}$.** Lines show the posterior mean ranking of each province's Omicron onset across 4 different $e_{min}$ values (based on 1.6 million posterior draws). On y axis, 1=earliest, 12=latest. Vertical bars show 95% credible intervals. The ranking pattern is stable across different values, with Noord-Holland consistently estimated as the first province experiencing Omicron onset.

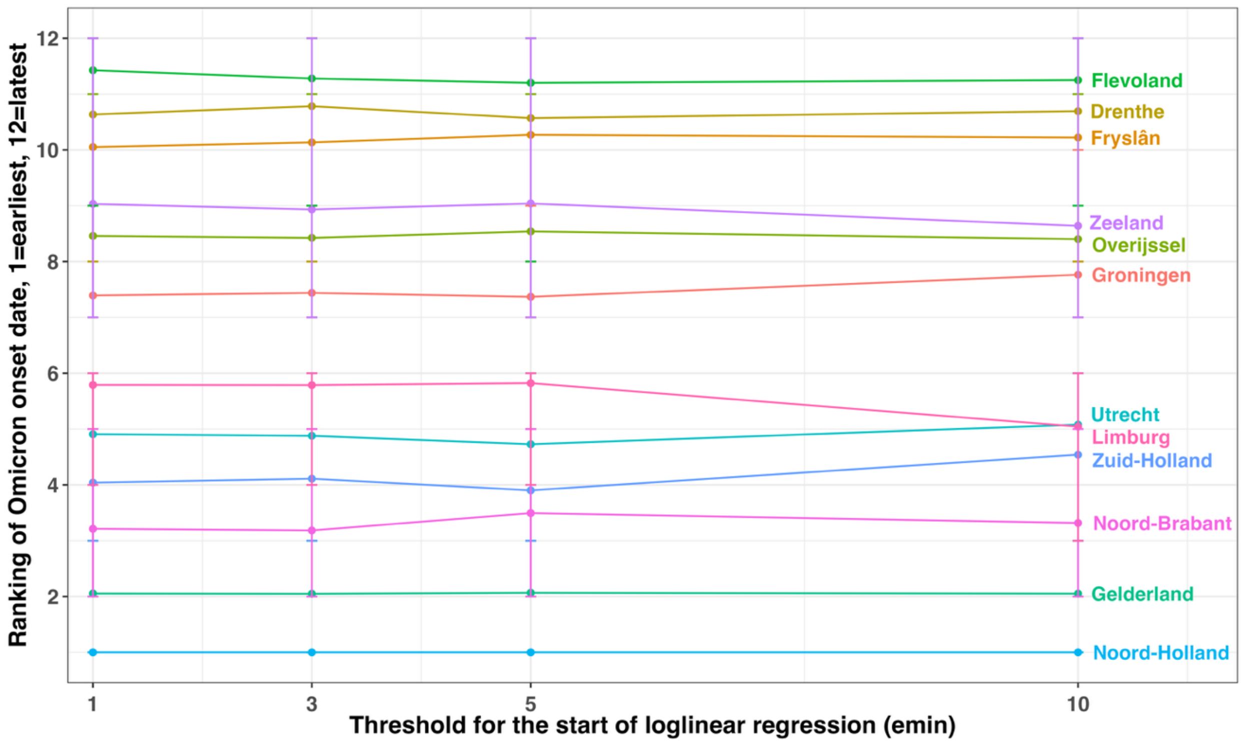

**Figure S4.2. Sensitivity of Omicron onset time ranking to the case threshold *I*.** Lines show the posterior mean ranking of each province's Omicron onset across six thresholds for weekly Omicron-positive new cases. On y axis, 1=earliest, 12=latest. Vertical bars show 95% credible intervals. The ranking pattern is stable for thresholds equal or larger than 30 cases per week, with Noord-Holland consistently estimated as the first province experiencing Omicron onset. Only a few provinces changed rank when thresholds are 10 and 20, largely reflecting greater stochastic variability when case counts are low. The default choice of $I = 30$ in the main analysis is a balance between the early-phase signalling and lower noise-driven mis-ranking.

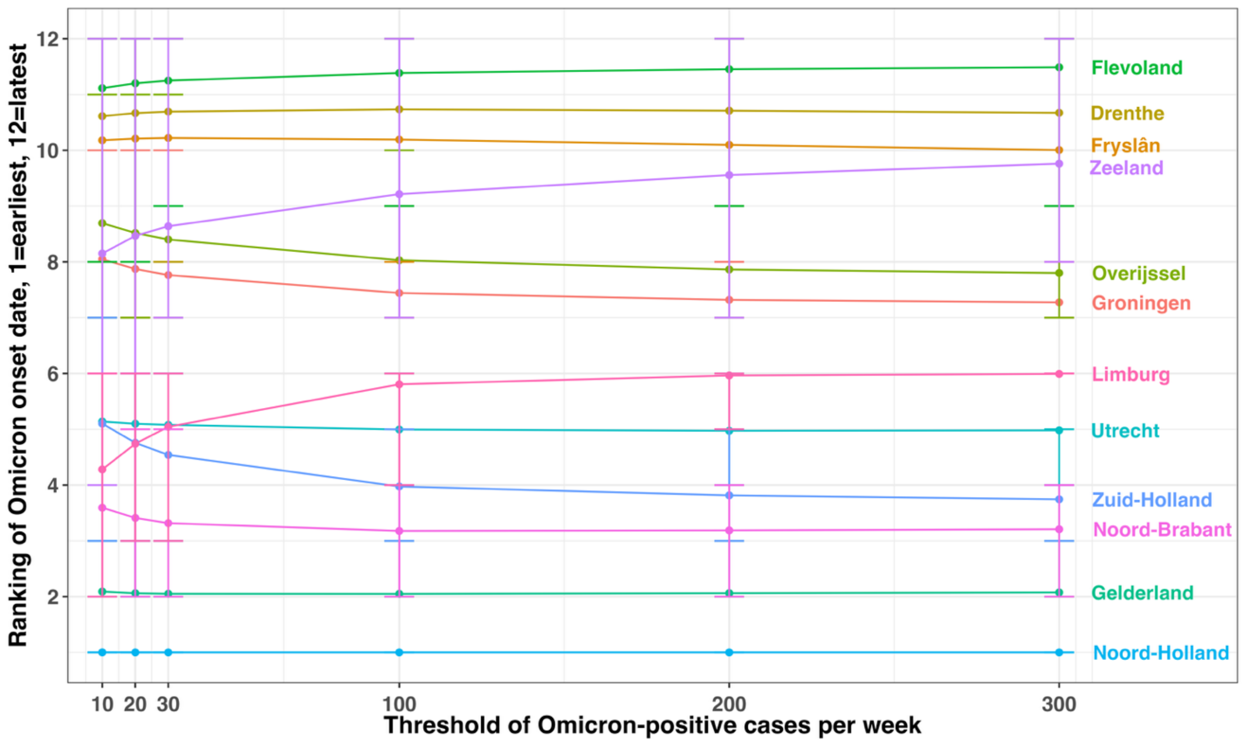

**Figure S5. Association between connectivity with Noord-Holland and Omicron onset time.** A negative association indicates provinces with more colleague connections with Noord-Holland are associated with earlier Omicron onset. A negative slope of -0.53 on the standardized natural logarithmic scale indicates that a tenfold increase in within-province colleague connections is associated with an Omicron onset that is, on average, 7 days earlier. Full names of provinces and the corresponding abbreviations are shown in supplementary Table S1.2.

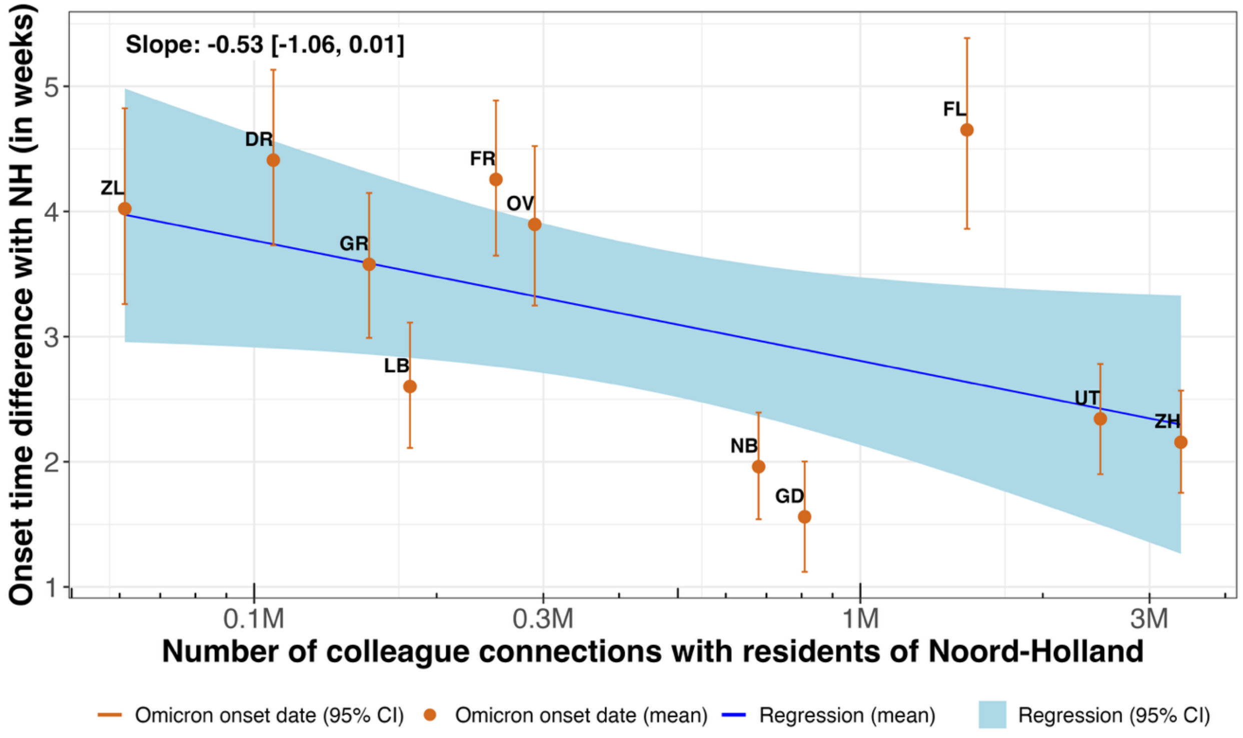

**Figure S6. Potential impact of safety-region-level lockdowns on colleague connections per residential safety-region pair.** Each panel visualizes the relative reduction in colleague connection between residential safety-region pairs when each of the 25 safety regions labelled at the top of the panel is placed under lockdown. Panel order is consistent with safety regions from north to south. The darker the shades, the larger proportions of connections will be removed.

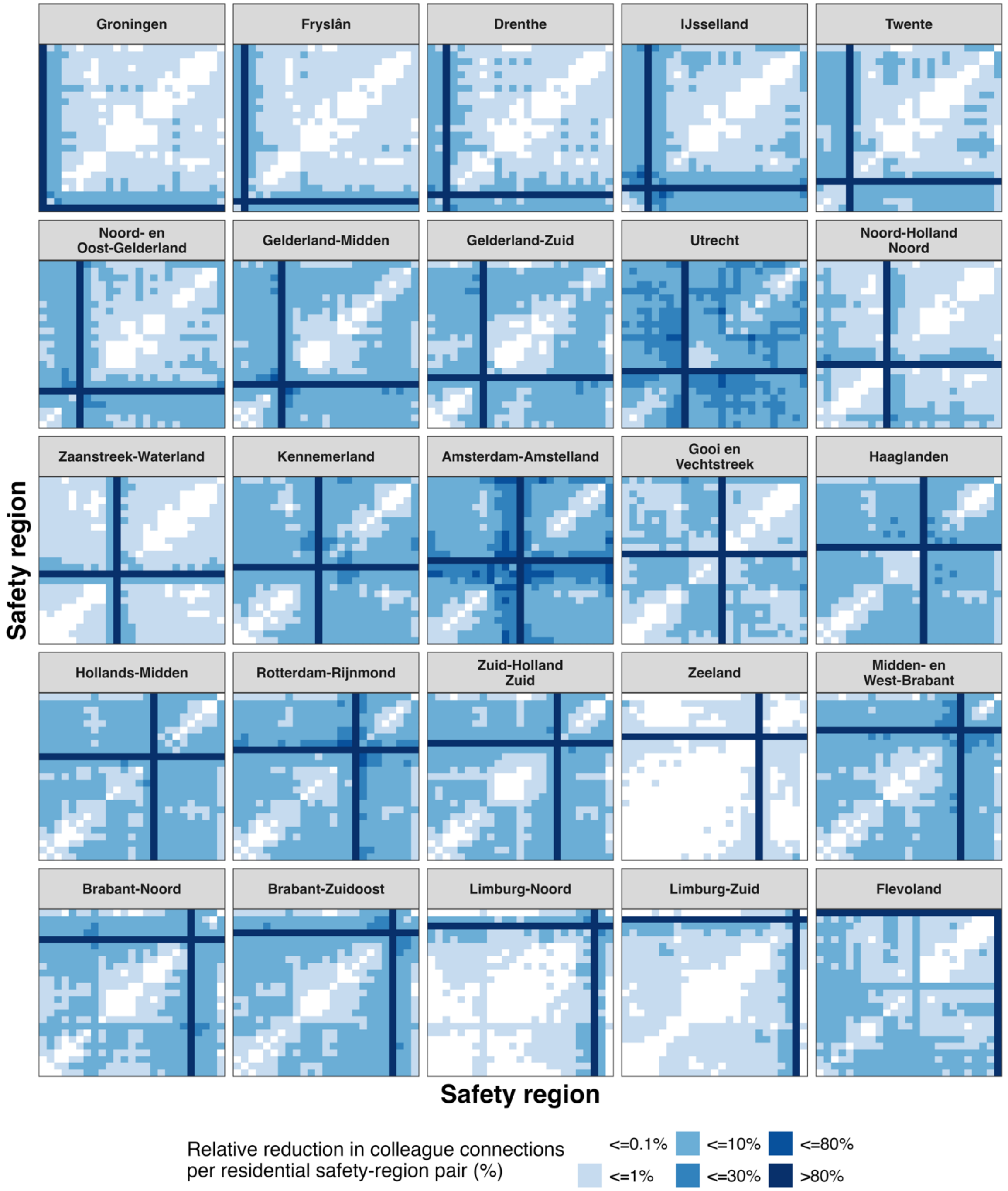

**Figure S7. Comparison of the potential impact of two regional interventions on total national colleague connections.** Fraction of total colleague links removed under a regional lockdown (x axis) compared to a cross-regional travel ban (y axis). Each circle indicates one of the 12 provinces (in blue), 25 safety regions (in orange) or 352 municipalities (in green). Grey dashed line indicates equal relative reductions under two regional interventions ($y = x$). Compared to regional lockdowns whose relative reductions on total national colleague connections are largely driven by regional labour force size, high populated regions are less dominant under cross-regional travel bans that remove only cross-regional links. A few example provinces and municipalities are labelled.

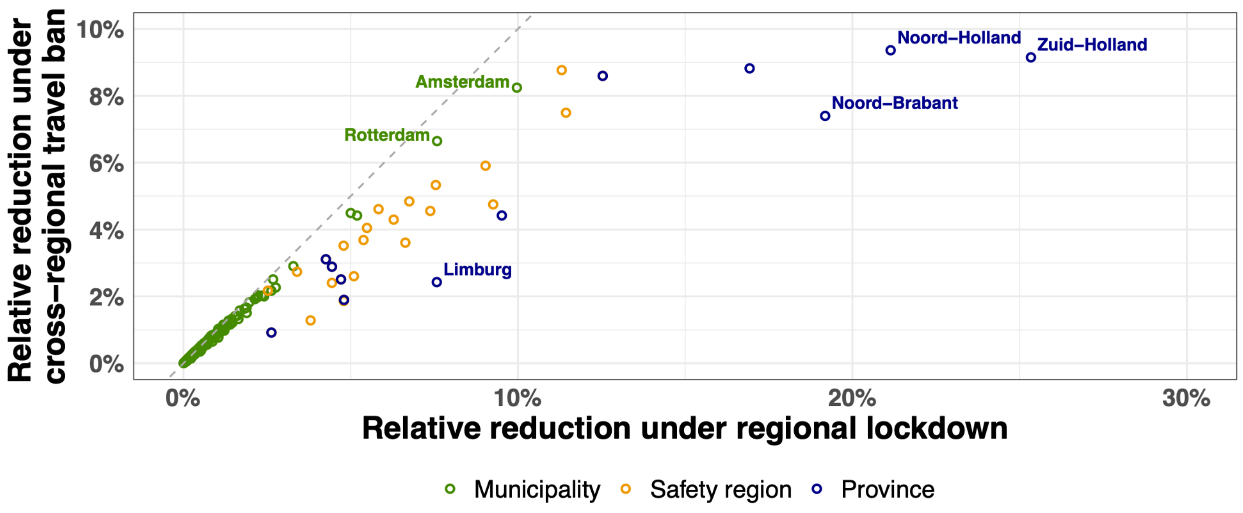
